# COVID-BioB Cohort Study: the neutralizing antibody response to SARS-CoV-2 in symptomatic COVID-19 is persistent and critical for virus control and survival

**DOI:** 10.1101/2021.02.17.21251929

**Authors:** Stefania Dispinseri, Massimiliano Secchi, Maria Franca Pirillo, Monica Tolazzi, Martina Borghi, Cristina Brigatti, Maria Laura De Angelis, Marco Baratella, Elena Bazzigaluppi, Giulietta Venturi, Francesca Sironi, Andrea Canitano, Ilaria Marzinotto, Cristina Tresoldi, Fabio Ciceri, Lorenzo Piemonti, Donatella Negri, Andrea Cara, Vito Lampasona, Gabriella Scarlatti

## Abstract

Understanding how antibody to SARS-CoV-2 evolve during infection may provide important insight into therapeutic approaches to prevent fatal COVID-19 illness and vaccines. Here, we profile the antibody response of 162 well-characterized COVID-19 symptomatic patients followed longitudinally for up to eight months from symptom onset. Using two newly developed assays we detect SARS-CoV-2 neutralization and antibodies binding to Spike antigens and nucleoprotein as well as to Spike S2 antigen of seasonal beta-coronaviruses, and to hemagglutinin of the H1N1 flu virus. Presence of neutralizing antibodies withing the first weeks from symptom onset correlates with time to a negative swab result (p=0.002) while lack of neutralization with an increased risk of a fatal disease outcome (HR 2.918, 95%CI 1.321-6.449; p=0.008). Neutralizing antibody titers progressively drop after 5-8 weeks but are still detectable up to 8 months in the majority of recovered patients regardless of age or co-morbidities. IgG to Spike antigens are the best correlate of neutralization. Antibody responses to seasonal coronaviruses are temporary boosted and parallel those to SARS-CoV-2 without dampening the specific response or worsening disease progression. Thus, a compromised immune response to the Spike rather than an enhanced one is a major trait of patients with critical conditions. Patients should be promptly identified and immediately start therapeutic interventions aimed at restoring their immunity.

## Introduction

The outcome of severe acute respiratory syndrome coronavirus 2 (SARS-CoV-2) infection while mild in the majority of infected individuals, is severe in a significant proportion, requiring hospitalization, and fatal in a minority.^1^

The SARS-CoV-2 antibody response has been studied in large cohorts of infected individuals with diverse Corona Virus Disease 19 (COVID-19) severity. Antibodies to the Spike’s receptor binding domain (RBD) appear within the first three weeks from symptoms onset and IgG and/or IgA seroconversion occur either sequentially or simultaneously with the appearance of IgM.^2,3^ Using a newly developed Luciferase Immunoprecipitation System (LIPS) assay to detect antibodies to SARS-CoV-2 antigens, we showed that 95% of 509 COVID-19 patients developed IgG by week four from symptom onset and further rose until the third month of follow-up.^4^ Also, the presence of RBD IgG in the earliest available sample correlated with survival while that of whole Spike IgA correlated with time to a virus negative naso-pharyngeal swab by RT-PCR.

The Spike glycoprotein mediates entry into target cells via the ACE2 receptor and neutralizing antibodies (nAbs) produced after infection in COVID-19 patients can block viral infection of human cells *in vitro* and thwart viral replication *in vivo*. Indeed, clinical trials with monoclonal nAb (mAb) against SARS-CoV-2 RBD, decreased viral load in patients with recently diagnosed mild/moderate COVID-19,^5–7^ suggesting that a native nAbs response could have a major impact on the disease if induced early after infection. However, there is still controversy on the impact of nAbs, with some early studies even suggesting a detrimental role on disease progression,^8,9^ while more recent data showed no differences in nAbs early after infection in hospitalized patients with diverse disease outcome.^3^ Moreover, data on the closely related MERS and SARS-CoV beta-coronaviruses suggest that humoral immunity, while still detectable beyond one year after hospitalization, wanes over time.^10,11^ Whether SARS-CoV-2 nAbs decline at a similar pace is yet to be established conclusively considering the current timing of the pandemic. As of today, most published serological studies are mainly cross-sectional or at the most include a longitudinal follow-up of few months.^3,12–14^

Here we provide a comprehensive antibody profile of well-characterized COVID-19 symptomatic patients followed longitudinally for up to eight months from symptom onset. We tested 162 patients using a newly developed, robust lentiviral vector (LV)-based SARS-CoV-2 neutralization assay and our LIPS assay for the detection of IgG, IgM and IgA to SARS-CoV-2 Spike, RBD and nucleoprotein (NP) as well as to Spike of seasonal beta-coronaviruses, and to H1N1 influenza virus hemagglutinin. Our results indicate that early development of nAbs is critical for patient survival and virus control, supporting the introduction of mAb therapy early on after infection in selected patients and that the induction of protective nAbs by prophylactic vaccination is likely durable.

## Methods

### Study population

We studied 162 patients with confirmed SARS-CoV-2 infection enrolled from February 25^th^, 2020 in the COVID-19 clinical-biological cohort study (COVID-BioB) at the IRCCS San Raffaele Hospital (ClinicalTrialsgov-NCT04318366) as previously described (Supplementary Materials, SM).^15^ Biological material for COVID-BioB included a dedicated serum sample collected at hospital attendance (in-hospital sample) either in the Emergency Department (ED) or the ward and follow-up samples collected in the out-patient clinic at planned 1, 3 and 6 months visits post-discharge. An in-hospital sample was available for 150 patients, while follow-up samples were available for 87 patients at the 1 month visit, 77 patients at the 3 month visit, and 46 patients at the 6 months visit (table 1). Twelve patients with no history of hospital admission were followed at the out-patient clinic for up to five visits.

**Table 1:**
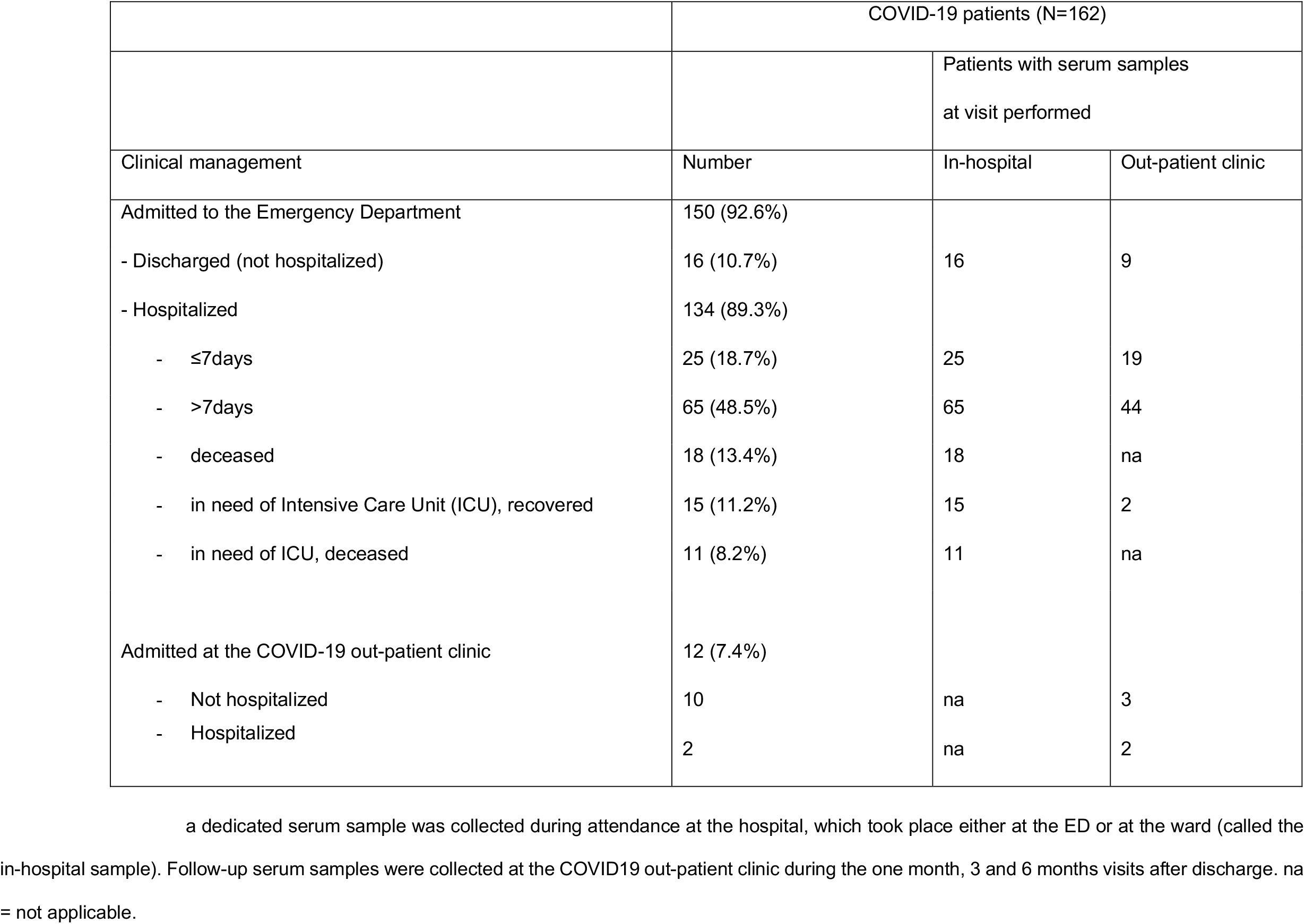
Clinical management and serum samples availability of the COVID-19 study patients.

|  | COVID-19 patients (N=162) |  |  |
| --- | --- | --- | --- |
|  |  | Patients with serum samples<br>at visit performed |  |
| Clinical management | Number | In-hospital | Out-patient clinic |
| Admitted to the Emergency Department | 150 (92.6%) |  |  |
| - Discharged (not hospitalized) | 16 (10.7%) | 16 | 9 |
| - Hospitalized | 134 (89.3%) |  |  |
| - ≤7days | 25 (18.7%) | 25 | 19 |
| - >7days | 65 (48.5%) | 65 | 44 |
| - deceased | 18 (13.4%) | 18 | na |
| - in need of Intensive Care Unit (ICU), recovered | 15 (11.2%) | 15 | 2 |
| - in need of ICU, deceased | 11 (8.2%) | 11 | na |
| Admitted at the COVID-19 out-patient clinic | 12 (7.4%) |  |  |
| - Not hospitalized | 10 | na | 3 |
| - Hospitalized | 2 | na | 2 |

### Fluid-phase luciferase immune precipitation (LIPS) assay

Antibodies were measured by LIPS assays.^4^ We expressed by transient transfection into Expi293F™ cells (Thermo Fisher Scientific, Waltham, MA, USA) several recombinant viral antigens tagged with a Nanoluciferase reporter (Promega, Madison, Wisconsin, USA) as described in SM and figure S2. Each antigen was incubated in liquid phase with test serum (1ul) for 2 h, followed by immune-complexes capture with rProtein A-sepharose and measurement of recovered luciferase activity in a XS960 Centro luminometer (Berthold). Using positive sera as standards, raw data was then converted into arbitrary units (AU).

### Lentiviral vector-based SARS-CoV-2 neutralization assay

LV expressing Luciferase (LV-Luc) pseudotyped with a membrane tethered Spike protein, either full length or with a 21 aminoacid truncated cytoplasmic tail (C3), or the VSV.G glycoprotein were produced by transient transfection of 293T Lenti-X cells with transfer vector plasmid pGAE-LucW, packaging plasmid pADSIV3+ and pseudotyping plasmid (pSpike, pSpike-C3 or phCMV-VSV.G) as detailed in SM. LV-Luc preparations were titered on VeroE6 cells and dilutions providing 150,000-200,000 RLU were used in the neutralization assay. Serum serial 3-fold dilutions starting from the 1/40 dilution were incubated in duplicate with the LV-Luc for 30 min at 37°C in 96-well plates, and thereafter added to VeroE6 cells. After 48 h luciferase expression was determined with a luciferase assay system (Bright-Glo, Promega) and measured in a Mitras luminometer (Berthold). Inhibitory serum dilution (ID) 50 was calculated with a linear interpolation method.^16^

### Statistical analysis

Median with either 95% CI, range, or inter-quartiles range (IQR) in parenthesis and frequency or percent were used to report continuous variables and categorical variables, respectively. Chi-square or Fischer’s exact test were used to compare categorical variables. Wilcoxon rank sum or Kruskal-Wallis test were used to compare continuous variables. Imputation for missing data was not performed. Time-to-event was calculated from the date of symptom onset to the date of death, or of last follow up visit, whichever occurred first. Survival was estimated according to Kaplan–Meier. Association between antibody positivity and time to death was calculated using univariate and multivariate Cox proportional hazards models. The effect was reported as hazard ratio (HR) with the corresponding 95% CI, estimated using the Wald approximation. All survival analysis and association were stratified according to time from the onset of symptoms to blood sampling (weeks 1, 2, 3, ≥ 4). Two-tailed p values are reported, with p value <0.05 indicating statistical significance. All confidence intervals are two-sided and not adjusted for multiple testing. Statistical analyses were performed with SPSS 24 (SPSS Inc./IBM) and the R software version 3.4.3.^17^

## Results

### COVID-BioB Study Cohort characteristics

We profiled the humoral immune response of 162 patients of our COVID-BioB cohort. All had a confirmed SARS-CoV-2 infection and a record of symptoms onset. Serum samples were collected at hospital admission in March-April and at the out-patient visits during follow-up until Nov 25, 2020 (table 1). Patients were predominantly Caucasian (89.5%), male (66.7%) and had a median age of 63 years. A relevant fraction had a body mass index >30 (32.4%) and 57.4% had one or more co-morbidity, with hypertension (44.4%) and diabetes (24.7%) being the most frequent (table 2). Patients sought an emergency hospital visit after a median of nine days from symptoms onset and 134 were hospitalized. At admission, symptoms consisted prevalently of fever, fatigue and respiratory difficulties often associated with dyspnea. The median hospitalization duration was 14 days. Overall, 26 patients entered the Intensive Care Unit (ICU) and 29 passed away, of whom 11 after ICU entry. The median time to a negative RT-PCR results of the naso-pharyngeal swab was 40 days (IC95% 37-43). Patients who recovered were followed-up for a median of 201 days (range 145-250) from symptom onset. The set of laboratory blood tests recorded as *per* COVID-BioB protocol is presented in table S1 and SM. The general characteristics of this study cohort reflect those of the overall COVID-19 patients admitted to our hospital during the same period and previously described.^4,15^

**Table 2:**
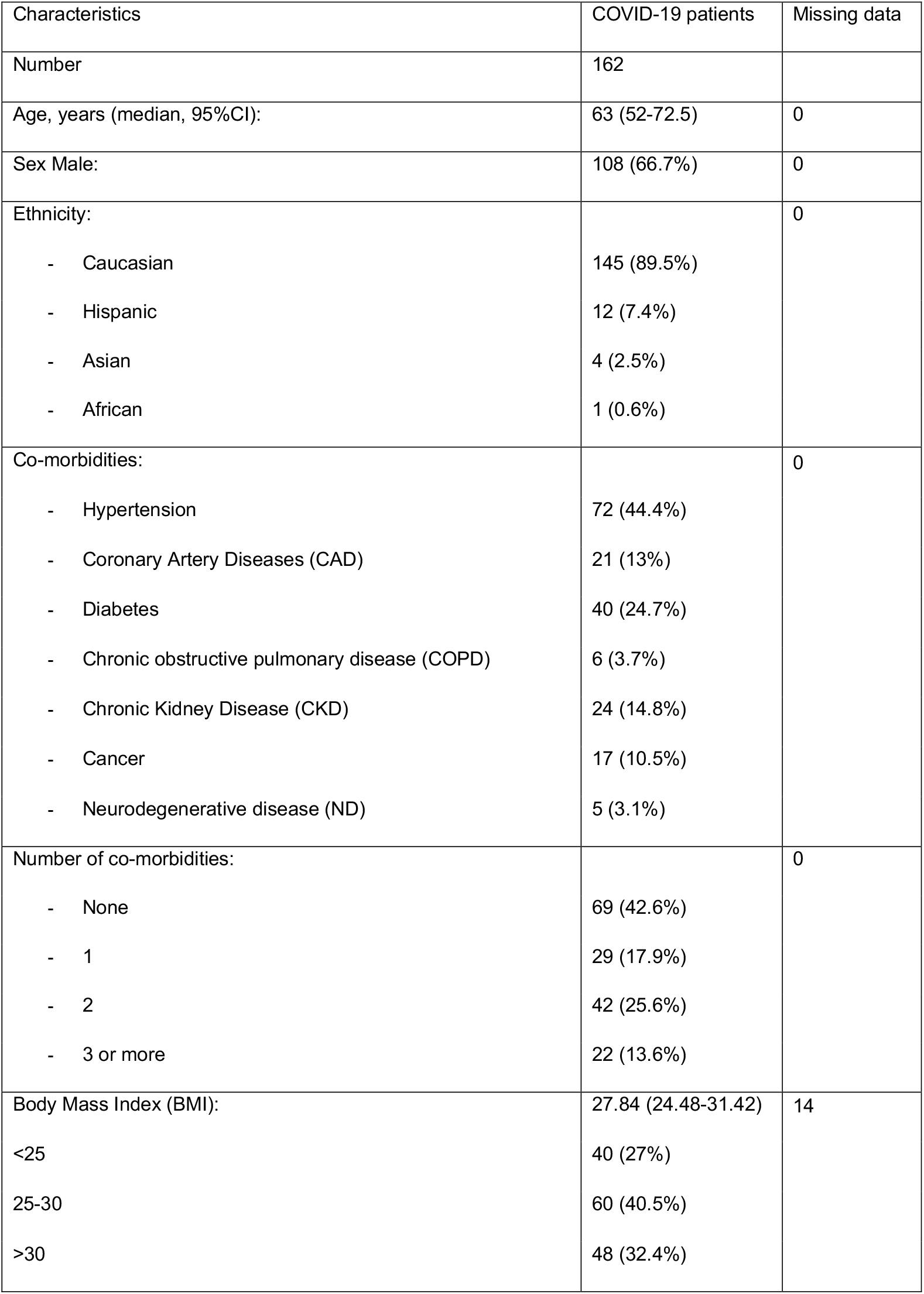
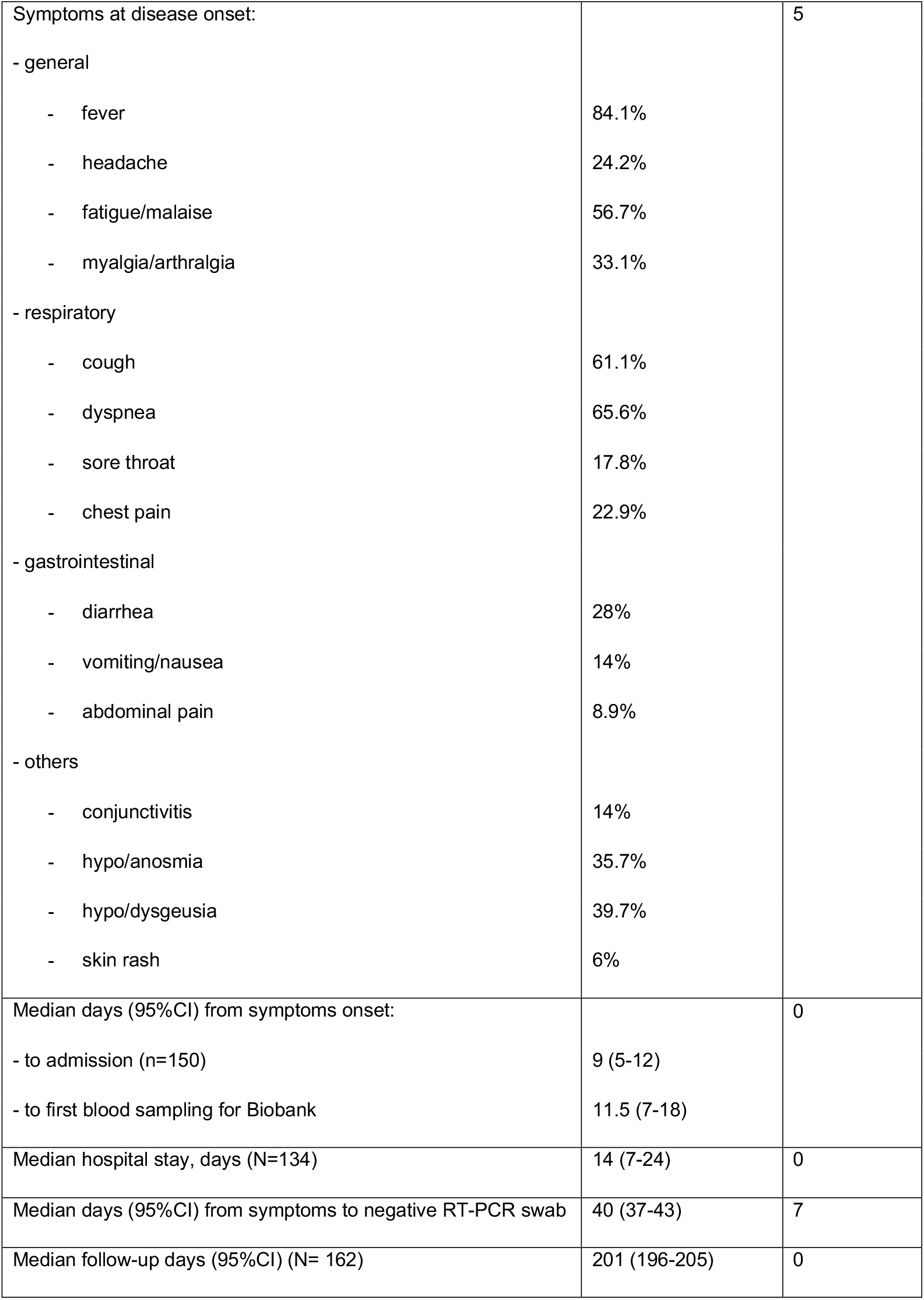
Characteristics of the COVID-19 study population.

### Truncated Spike on LV pseudovirions improves infectivity and is suitable for SARS-CoV-2 neutralization assay

To test the nAb response, we developed and optimized a Lentiviral (LV)-based SARS-CoV-2 neutralization assay, suitable for handling in a Biosafety Level 2 environment. This assay is based on Luciferase expressing LV (LV-Luc) pseudoparticles, enveloped with a Spike glycoprotein containing a 21aa deletion in the cytoplasmic tail (Spike-C3) (SM, figure S1). This truncation removes an endoplasmic reticulum (ER) retention signal that interferes with the budding of viral particles,^18,19^ thus favoring the interaction of Spike and Gag proteins during particle assembly.^20^ Upon transfection into 293T Lenti-X cells of either full-length Spike or Spike-C3 expression plasmids, both constructs showed comparable expression levels on the cell surface (figure 1A). Conversely Spike-C3 was incorporated on the pseudoparticles’ surface at higher levels than the whole Spike, though Gag protein content was comparable across all preparations (figure 1B).

**Figure 1:**
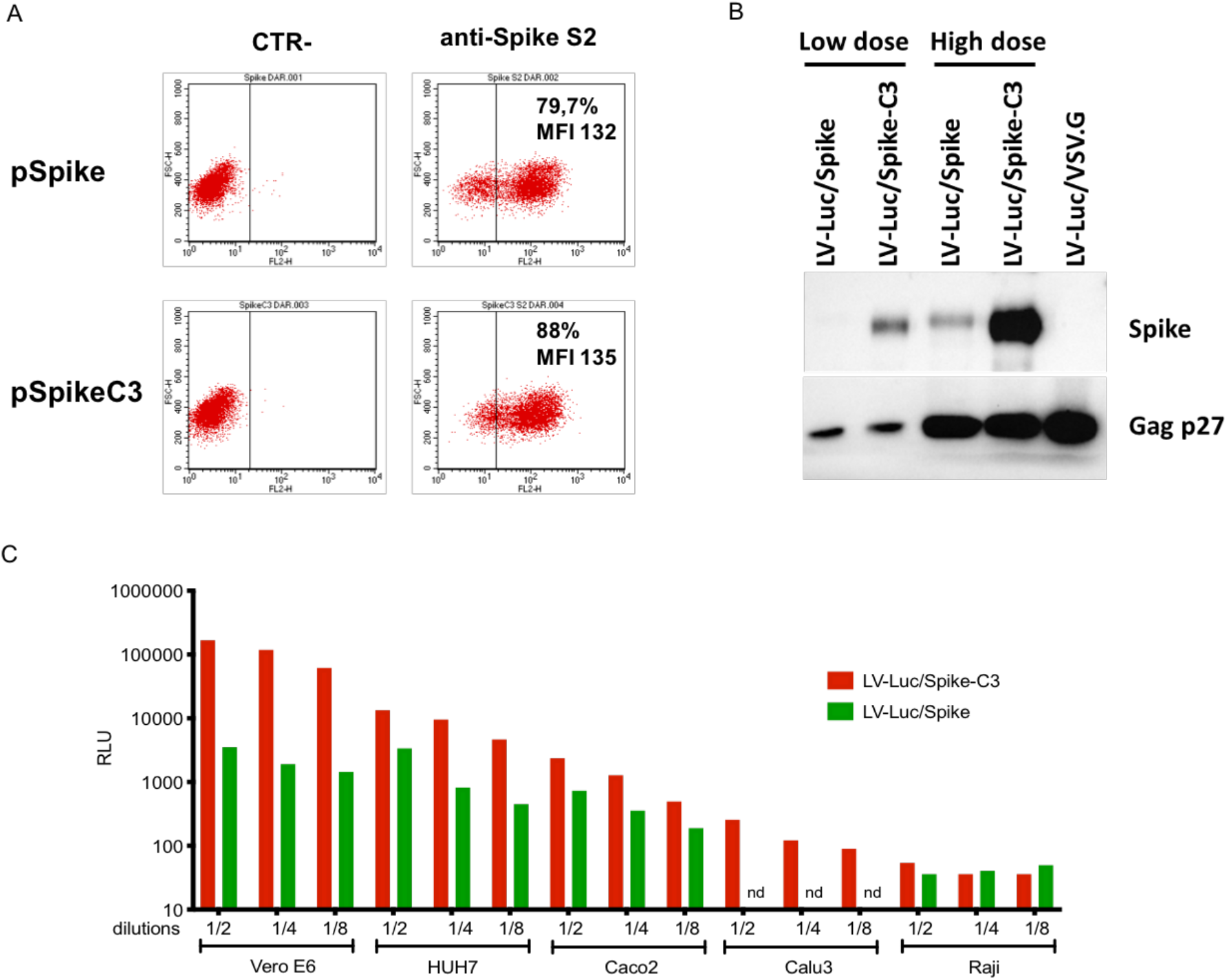
Higher incorporation of truncated Spike on LV pseudovirions improves infectivity in ACE2+ cell lines. (A) Evaluation of SARS-CoV-2 Spike expression in 293T Lenti-X cells transfected with pSpike and pSpike-C3 plasmid by flow cytometry. (B) Western blot (WB) of lysates from concentrated preparations of LV-Luc pseudotyped with wild type Spike (LV-Luc/Spike) or Spike truncated (21aa) at the cytoplasmic tail (LV-Luc/Spike-C3). Low dose: 1×10^5^ RT units; High dose: 6×10^5^ reverse transcriptase (RT) Units. Control vector included LV-Luc pseudotyped with VSV.G envelope (LV-Luc/VSV.G). (C) Infection of human and macaque epithelial cell lines and the control B cell line Raji incubated with decreasing concentrations of LV-Luc pseudotyping the full length (LV-Luc/Spike) or the truncated form (LV-Luc/Spike-C3) of the SARS-CoV-2 Spike. Data are expressed as mean luciferase activity (RLU, Relative Light Units) of duplicates.

LV-Luc/Spike and LV-Luc/Spike-C3 infectivity was tested in a panel of human and macaque epithelial cell lines. Consistently with the higher incorporation of Spike-C3 on the pseudoparticle surface, LV-Luc/Spike-C3 was always more infectious than LV-Luc/Spike, with an infection capacity three to 50 times higher depending on the cell line used (figure 1, SM). For further assay development we selected a combination of LV-Luc/Spike-C3 construct and VeroE6 monkey epithelial kidney cell line that showed the highest ACE2 expression (data not shown). The optimization of key assay parameters is described in SM.

### Antibody responses to SARS-CoV-2 Spike appear early after symptoms onset though not in the totality of patients

Using our two newly developed assays we profiled the sera of 162 COVID-19 patients for the SARS-CoV-2 nAb response and for the antibody binding to SARS-CoV-2 Spike domains (S1+S2, S2, RBD) and NP, the Spike S2 of HCoV-OC43 and HCoV-HKU1 and the HA1 protein of the 2009 H1N1 flu virus. Sera were available for 150 patients during the hospital stay, with the sampling occurring between 1-38 days from symptom onset. Early nAb responses showed a large heterogeneity in magnitude (median ID50 1/786, range 1/<40-1/1,660,000) (figure 2A, table S2). nAbs were present in 43.2% patients (19/44, median ID50 1/410) sampled within the first week and in 78.9% (45/57, median ID50 1/1165) within the second week from symptom onset, respectively. Overall, 28.7% (43/150) of patients had no detectable nAbs in their in-hospital sample. IgM and IgA binding to the Spike proteins were more frequently detected than IgG during the first two weeks from symptoms onset (figure 3A). Spike and NP antibodies showed similar frequencies in samples taken 3-4 weeks post-symptom onset. The best binding antibody correlate of nAbs were IgG to any of the Spike antigens used in LIPS (figures 3B-C, S3), Spike IgG thus behaving as a good surrogate marker for nAbs detected in our neutralization assay.

**Figure 2:**
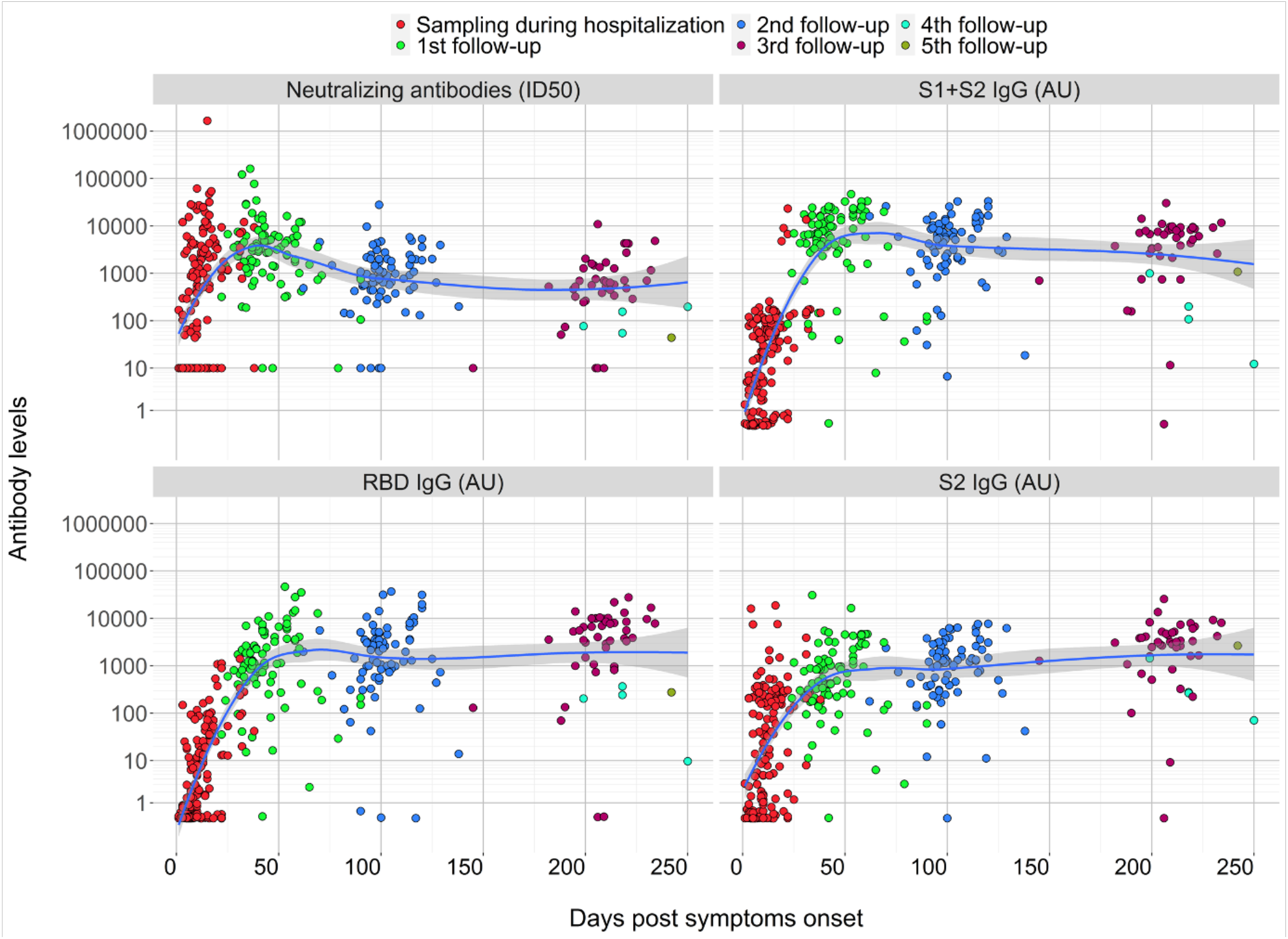
Kinetics of the anti-Spike antibody response. Scatterplots of each patient anti-Spike neutralizing, S1+S2 IgG, RDB IgG, S2 IgG antibodies over time from symptoms onset. Antibody levels correspond to the reciprocal of the ID50 for nAbs or to arbitrary units for all other reactivities. Sampling during hospital attendance (ER or ward) or at post discharge outpatient follow-up visits is shown by the indicated color code. Shown is the moving average of data + SE (black curve line + grey band) as obtained by a LOESS curve fitting polynomial regression.

**Figure 3:**
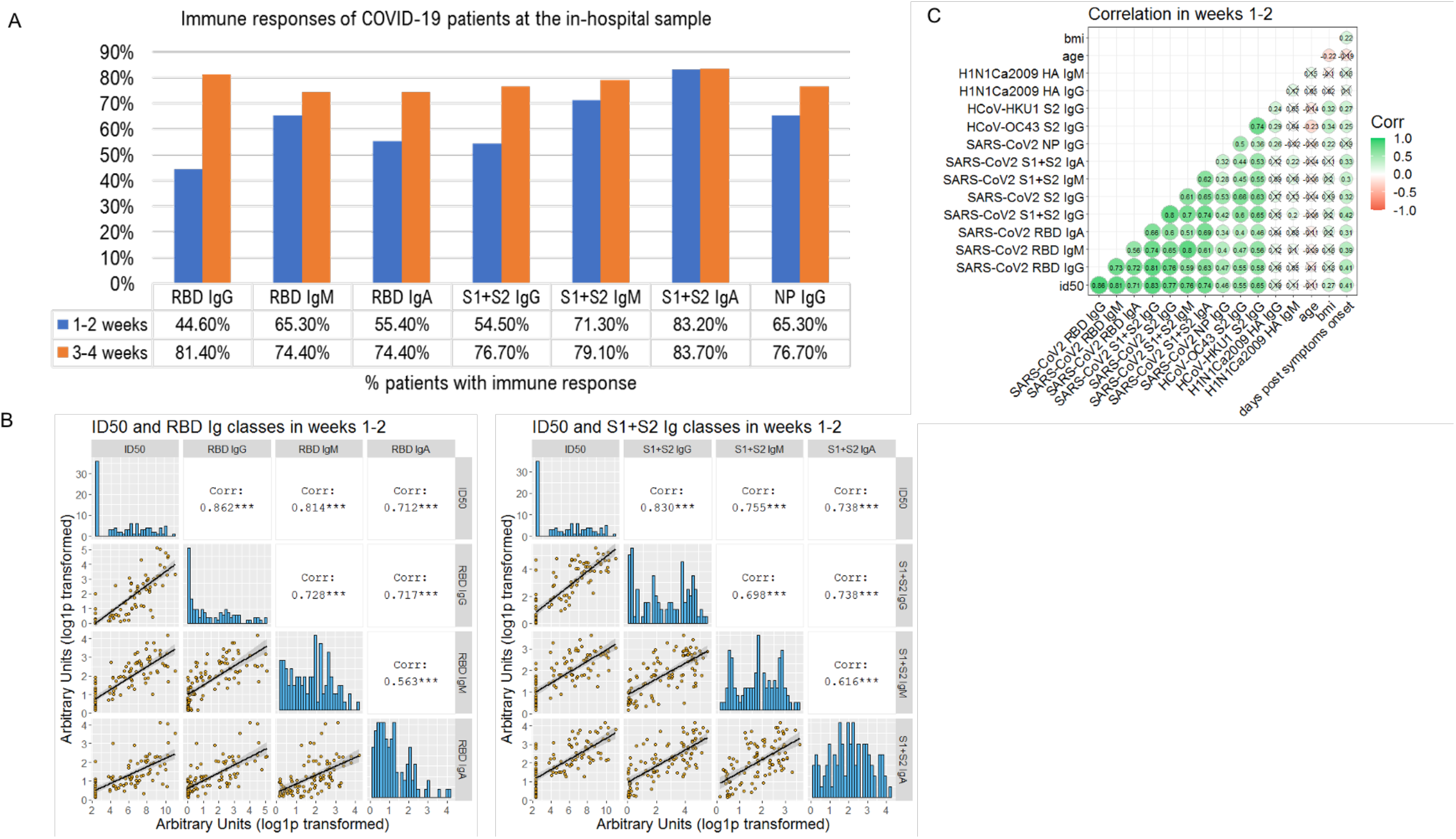
Anti-SARS-CoV-2 Spike neutralizing and binding antibody profile. (A) Indicated is the percent of 150 COVID-19 patients tested at the in-hospital visit, that had IgG, IgM and IgA to SARS-CoV-2 antigens detected with LIPS, when sampled during the first or second two weeks from symptoms onset. (B) Sera of COVID-19 patients, collected at the indicated timepoints from symptoms onset, were measured by LV based-neutralization assay and the LIPS indicated in grey labels above each row/column. Boxes under the diagonal show each correlation plot of the reciprocal of ID50 and arbitrary units after log10 conversion. Dots correspond to individual measurements, the black line represents the regression line and the grey area its 95%CI. Boxes on the diagonal show as histograms the distribution of values in each assay. Boxes above the diagonal show the corresponding Pearson correlation analysis coefficients. (C) Correlation matrix of the indicated variables in week 1-2. For each pair, the Pearson’s correlation coefficient is shown as number and on a color scale. Statistically non-significant correlations are crossed.

### Kinetics and persistence of antibodies to SARS-CoV-2

The longitudinal follow-up showed that at the first out-patient’s visit (median 42 days from symptoms onset, range 22-90), nAbs were still rising in 71.6% of the 87 patients sampled (median ID50 1/3190; range 1/<40-161,000). While at the second visit (median 99 days, range 62-138) nAbs declined in all 77 tested patients (median ID50 1/909, range 1/<40-28,000) (figure 2A, table S2), with a complete loss of nAbs in just three patients. Forty-six patients attended additional visits at or beyond 6 months (median 204 days, range 145-250). In these patients nAb titers were further decreasing (median ID50 1/660, range 1/<40-10,900), but no additional patient lost the nAb response. In 26 non-hospitalized patients, despite a less severe disease, the profile of nAbs and anti-Spike antibodies was similar to that of hospitalized patients (figure S8, SM). Overall, 67.3% (101/150) COVID-19 patients developed nAbs rapidly within 2 weeks from symptoms onset, which persisted up to 36 weeks in all but three patients. As expected, IgM and IgA to SARS-CoV-2 antigens, measured up to eight weeks from symptoms onset, progressively declined during follow-up (figures 3B-C). The kinetic of IgG response to Spike proteins reflected that of nAbs (figure 2, table S2): antibodies to S1+S2 reached a peak by 5-8 weeks and then declined, while those to RBD and S2 continued to rise throughout follow-up, but still correlated significantly with nAbs (figures 3, S3).

### Development of neutralizing antibody after symptoms onset correlates with viral control

Prevention of a critical COVID-19 illness is a major hurdle and biomarkers are strongly needed to drive the choice of therapeutic interventions. We performed an extensive analysis of antibody responses and clinical and laboratory parameters collected at or in close proximity of the in-hospital sampling in relation to time to a negative RT-PCR result of the naso-pharyngeal swab and to patient’s survival. In the univariate analysis the presence of nAbs was the only other positive strong predictor of time to a negative swab test (HR 2.238; 95%IC 1.343-3.734) besides IgA to SARS-CoV-2 S1+S2 (HR 1.958; 95%IC 1.079-3.554) (figure S4). Early development of nAbs significantly correlated with the time to a negative swab (p=0.002), i.e. clearance of the virus, when stratified for time from symptoms onset (figure 4A). It was instead impossible to investigate the correlation with viral load at disease onset since a quantitative RT-PCR assay was not yet available when swabs were analysed.

**Figure 4:**
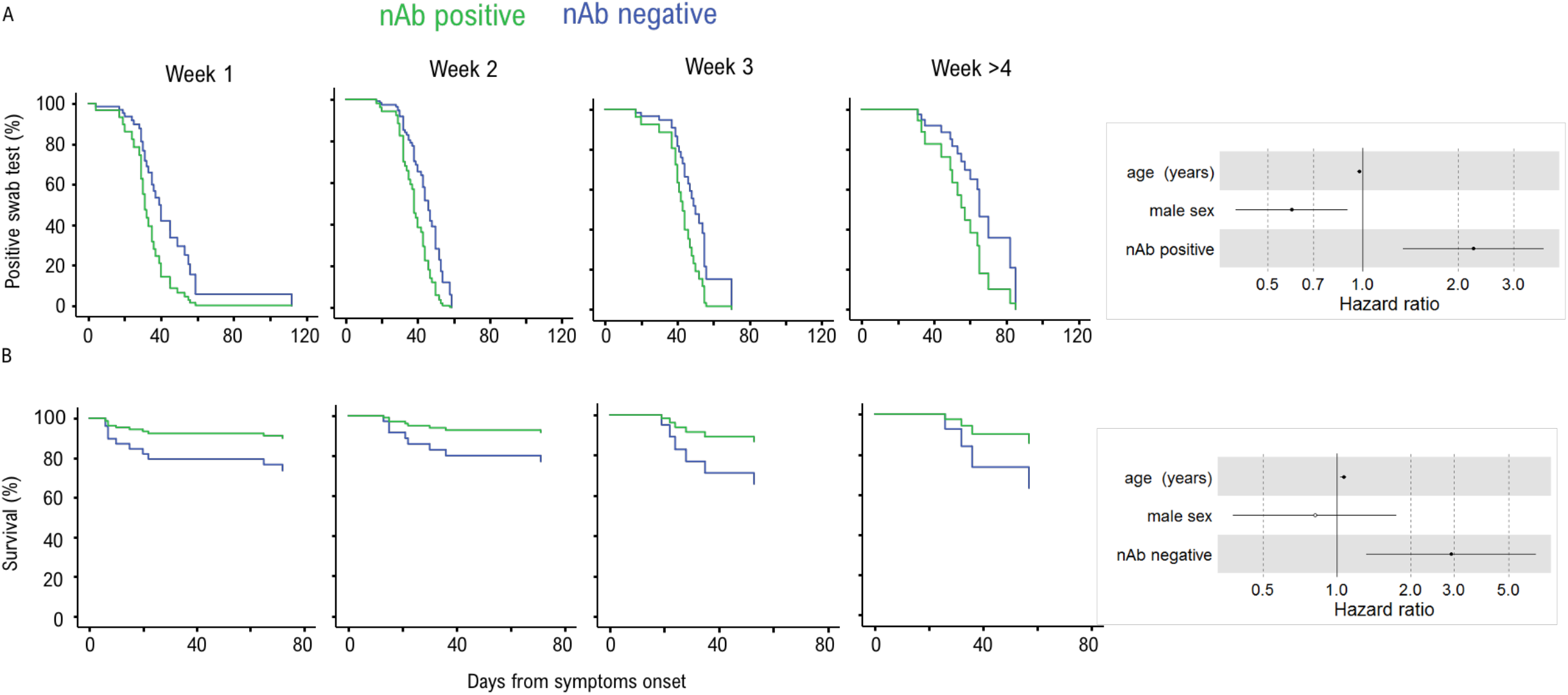
Early development of the nAb response correlates with viral control and survival. Cox regression *s*urvival estimates for 150 patients with COVID-19 sampled for nAb response to SARS-CoV-2 during in-hospital visit. The analysis was adjusted for sex and age and stratified for the time from symptoms onset at serum sampling. In (A) the time to a SARS-CoV-2 negative naso-pharyngeal swab (HR 2.238, 95%CI 1.342-3.734; p=0.002) and in (B) the survival rate (HR 2.918, 95%CI 1.321-6.449; p=0.008) was estimated by the presence or absence of a nAb response. Dots represent the Hazard Ratio (HR), filled dots stand for p<0.05.

### Lack of a neutralizing antibody response predicts fatal outcome

If developed early after infection, nAbs may limit viral spread, and consequent disease progression. In the 134 hospitalized patients, nAbs detected at the time of the in-hospital sampling did not show any impact on the hospitalization length (table S3), while significantly correlated with critical illness (figure S4). Importantly, lack of a nAb response early after infection was a strong predictor of death when stratified for time from symptoms and corrected for age and sex (HR 2.918, 95%CI 1.321-6.449; p=0.008) (figure 4B). This correlation was still maintained if we applied more stringent ID75 (HR=2.67 (1.2-5.9), p=0.015). In addition, the correlation was further maintained when corrected for each one of the identified correlates of death, including co-morbidities and relevant immune-inflammatory markers, and the absence of IgA to S1+S2 (figure S5).

### Kinetics and persistence of neutralizing antibodies is preserved despite advanced age and co-morbidities

We investigated also the impact of advanced age and diverse co-morbidities on nAb response’ kinetics (table S3). In patients above 63 years of age or presenting one or more co-morbidities and who passed away, we detected low or no nAbs in the first weeks after symptoms onset (figure 5). Patients without an early nAb response were significantly older with debilitating co-morbidities, including cancer or chronic pulmonary and kidney diseases. Noteworthy, these patients presented frequently a disease onset with less severe COVID-related respiratory symptoms that did not require ICU admission, and clinical laboratory tests mostly showing a lower immune-inflammatory activation with organ involvement. Conversely, in patients who recovered, advanced age and presence of co-morbidities did not impact on the distribution of early antibody response and, analogously, titre, kinetics and persistence of nAbs after recovery were not affected by either parameter.

**Figure 5:**
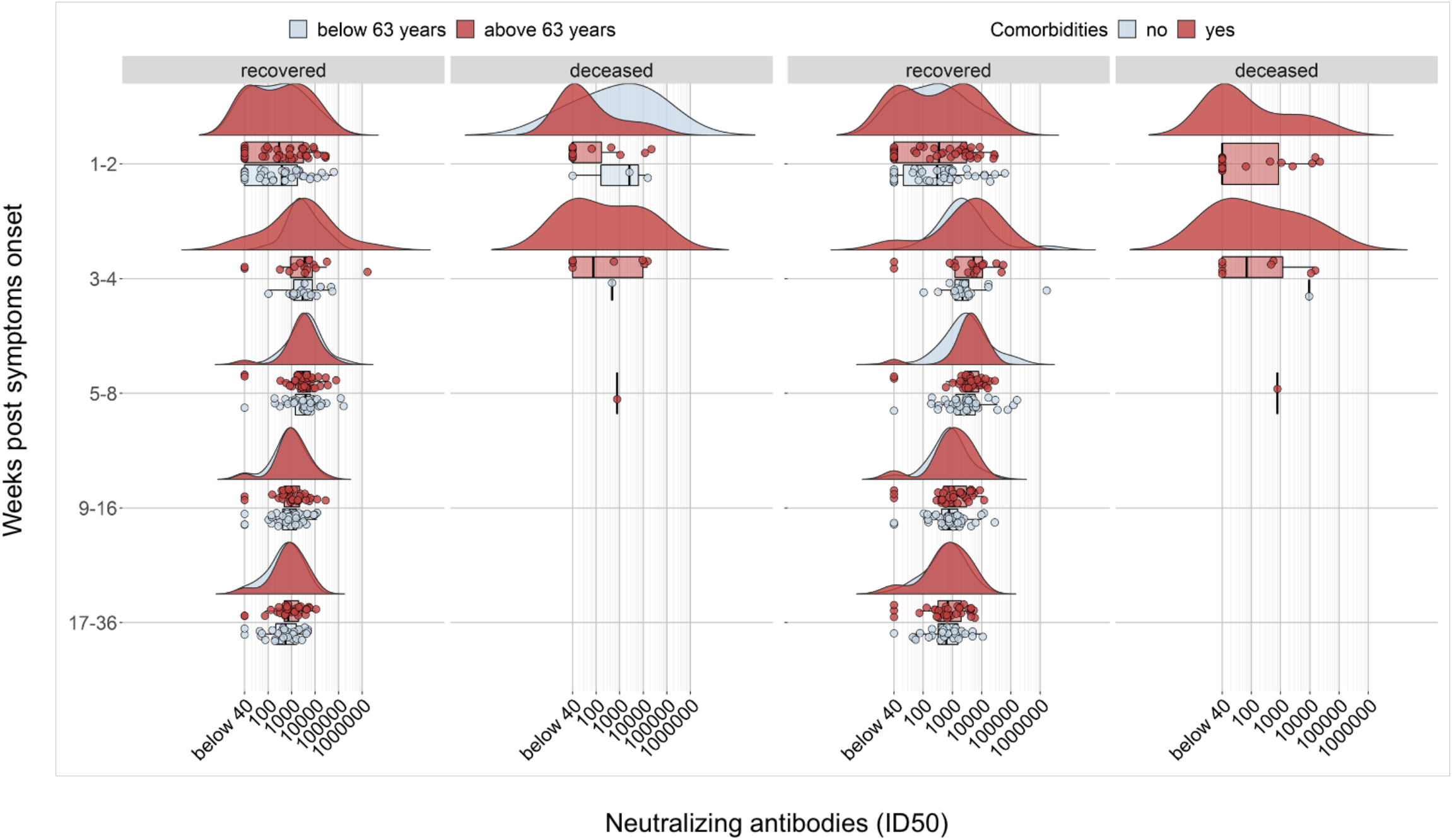
Development and kinetics of the nAb response according to age or co-morbidities. Raincloud plot of each patient’s nAb response (circles) expressed as the reciprocal of the ID50 titer. Patients are stratified by time from symptoms onset at sampling (vertical panels, weeks) and clinical outcome (recovered or deceased). Patients are further stratified as below or above the median age of the overall study population (<63 years = blue, >63 years = red) or according to the presence of any co-morbidity (present = red, absent = blue). Shown are the probability density estimate (with the half violin plot upscaled to maximum width for better visualization) and box plots displaying median, IQR, and whiskers extending to 1.96 times the IQR.

### Antibody responses to seasonal coronaviruses are temporary boosted without worsening disease progression

Pre-existing antibody responses to other beta-coronaviruses have been proposed to potentially have either a detrimental or beneficial role in COVID-19.^21,22^ Testing of IgG to S2 of HCoV-OC43 and -HKU1 in samples collected during the first weeks from symptoms onset did not correlate with either time to a negative swab test or death (figure S4). However, in the same time window, IgG titers to S2 of HKU1 and OC43 correlated with SARS-CoV-2 nAb titers (figures 6A, S6). Interestingly, patients who developed nAbs to SARS-CoV-2 during the first three weeks from symptoms onset had significantly higher IgG titers to either HCoVs compared to patients with undetectable nAbs, a difference that persisted throughout follow-up (figures 6B-C). Conversely, a pre-existing or ongoing Flu infection as documented by an antibody (IgM and/or IgG) response to the H1N1 flu virus HA antigen in the in-hospital samples did not correlate with presence of SARS-CoV-2 nAbs nor survival (figure 3B, table S2), further indicating that the immune response was not compromised in general.

**Figure 6:**
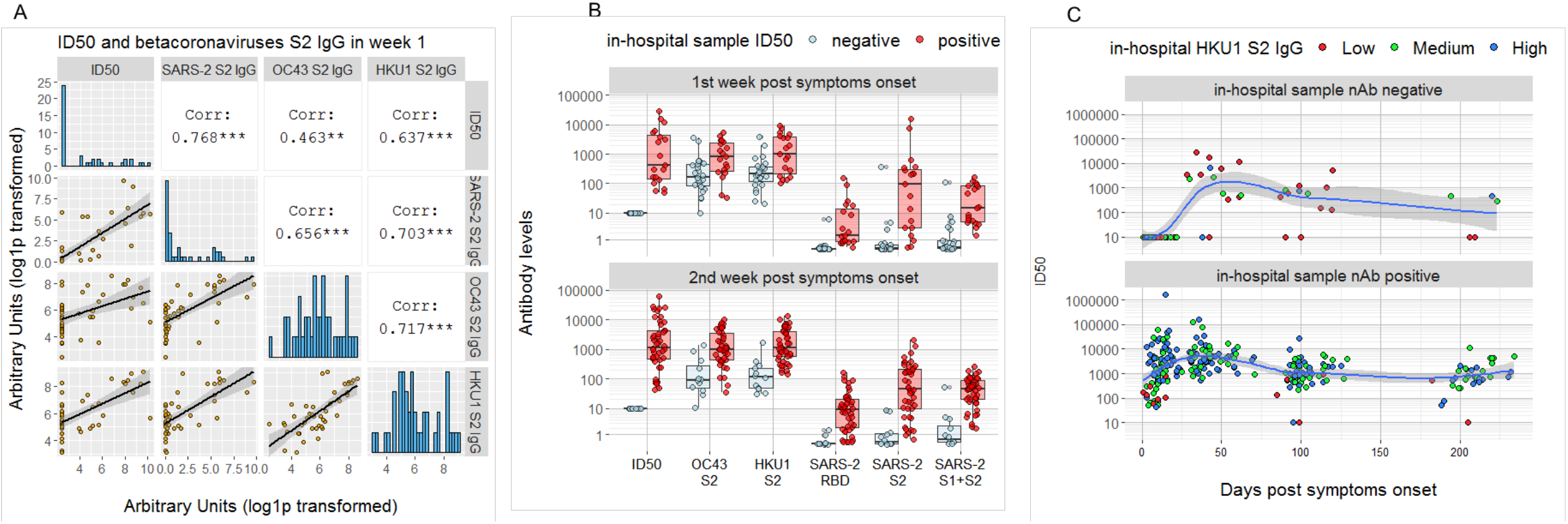
NAbs and antibody (IgG) to S2 HCOV-OC43 and HKU1 and SARS-CoV-2. Sera of COVID-19 patients, collected at the indicated timepoint from symptoms onset, were measured by LV based-neutralization assay and the LIPS to SARS-CoV-2 and HCoV S2 Spike antigen as indicated. (A) Boxes under the diagonal show each correlation plot of the reciprocal of the ID50 and arbitrary units after log1p transformation. Dots correspond to individual measurements, the black line represents the regression line and the grey area its 95%CI. Boxes on the diagonal show as histograms the distribution of values in each assay. Boxes above the diagonal show the corresponding Pearson correlation analysis coefficients. (B) Shown are box plot displaying median of each antibody response as indicated, IQR, and whiskers extending to 1.96 times the IQR, stratified by sampling at the indicated week from symptoms onset and a negative or positive nAb response. (C) Development and kinetics of nAb responses to SARS-CoV-2 during follow-up of patients with negative or positive nAb score at the in-hospital sampling. Each coloured dot corresponds to the reciprocal of the ID50 of a serum of a given infected individual stratified for low, medium or high IgG to HCoV S2 antigens. Shown is the moving average of data + SE (black curve line + grey band) as obtained by a LOESS curve fitting polynomial regression.

## Discussion

We used two newly developed and robust assays to profile comprehensively the longitudinal nAb response in symptomatic COVID-19 patients infected during the first wave of the pandemic. Our study contributes to the understanding of three important aspects of the SARS-CoV-2 infection, with relevant implications for the future of the pandemic. Firstly, nAbs are a correlate of survival as well as of virus control; secondly, nAbs and anti-Spike IgG persist in the vast majority of recovered patients regardless of disease severity, age and co-morbidities for up to eight months from symptoms onset; thirdly, SARS-CoV-2 infection back-boosts pre-existing antibody responses to seasonal beta-coronaviruses without dampening the development of neutralizing or binding SARS-CoV-2 antibodies nor enhancing critical illness.

Recently, Zohar et al. reported the correlation of critical disease outcome with lack of specific functional IgG responses but not with nAbs in COVID-19 patients.^3^ Previous reports raised doubts as to the efficacy of the protection conferred by nAbs in severe COVID-19 and suggested that enhanced nAbs might correlate with a negative clinical outcome.^12–14^ Conversely, in our study an absent nAb response early after disease onset, more than a difference in titre, was the strongest correlate with both death and delayed viral control. Aware that our patient cohort was characterized by an advanced age and diverse co-morbidities, we evaluated their impact on nAb kinetics. Indeed, the patients that did not develop nAbs were mostly elderly men (median age 74 years, range 59-79) with chronic severe co-morbidities. However, upon controlling for these factors, we confirmed that a compromised immune response to the Spike (rather than an enhanced one) is a major trait of patients with critical conditions. Conversely, patients with nAbs and anti-Spike IgA showed a faster control of the virus. Importantly, anti-Spike binding antibodies correlated with nAbs, confirming their potential usefulness as proxy biomarkers of a nAb response.

Recent data suggest that a cocktail of virus-specific mAbs was more efficient in reducing the viral load in symptomatic out-patients with high viral load but as yet no IgG and IgA response.^5^ Similarly, plasma with high titer nAbs administered to elderly patients within 3 days after symptoms onset reduced the risk of progression to severe respiratory disease.^6^ Currently, the Food and Drug Administration has granted an emergency use authorization for the virus-specific mAb therapy only for out-patients with mild-moderate COVID-19.^23^ Considering that elderly males with chronic co-morbidities are extremely vulnerable to severe COVID-19 in the absence of an anti-Spike response, we believe that these patients should be promptly identified and immediately start therapeutic interventions aimed at restoring the lack of a functional humoral immunity.

Interestingly, our data show that a pre-existing immunity to seasonal HCoVs may favour early and increased responses to SARS-CoV-2. In contrast with Aydillo et al. and in agreement with Anderson et al.,^21,22^ that analyzed smaller cohorts over a shorter time span, we found no correlation between the expansion of IgG to HCoVs and a delayed development of SARS-CoV-2 specific antibodies. In fact, our patients exhibited a back-boosting of antibodies to OC43 and HKU1 S2. This is hardly surprising in light of the structural and partial sequence homology (approximately 40%) between SARS-CoV-2 and HCoVs S2 domains. Indeed, the correlation of nAb titers with the IgG responses to seasonal beta-coronaviruses was good in the first weeks from disease onset, although invariably less than that with SARS-CoV-2 Spike IgG. However, with our LV-based assay we excluded that HCoV-S2 IgG positive, but SARS-CoV-2 Spike IgG negative sera neutralize SARS-CoV-2. Thus, if IgG to HCoV S2 are indeed cross-reactive to SARS-CoV-2 they are likely of low affinity and non-neutralizing but may possibly mediate a phenomenon of antibody dependent enhancement. This suggests that a pre-existing strong immunity to other beta-coronaviruses may in fact favour, more than deviate, an early nAb response to SARS-CoV-2.

As generally expected after the resolution of a viral infection, in our study nAbs started to drop in recovered patients after 5-8 weeks from symptoms onset. Nevertheless, in all but three patients nAbs above the assay threshold persisted for as long as eight months. Neither COVID-19 severity at disease onset, nor age, nor the presence of multiple co-morbidities affected the kinetics and persistence of nAbs. At the last follow-up visit, the distribution of nAbs titre was wide and spanning four orders of magnitude starting from close to the assay threshold. However, regardless of presence of long-term detectable antibodies, infection and vaccination will induce a memory B-cell response,^14,24^ that might rapidly mount a specific humoral response. At the third follow-up visit in nine of the 42 nAb-positive COVID-19 patients we documented a major increase of nAb titre (up to 400%) paralleled by levels of IgG to Spike antigens (figure S7). We cannot conclusively state that this raise was due to a re-exposure to SARS-CoV-2 since none of the patient declared any COVID-related symptoms nor was a virus detection test performed. However, the time of sampling during follow-up of these patients overlapped with the second epidemic wave in Italy.^25^ We can speculate that if indeed a re-exposure to the virus occurred, followed by either an asymptomatic or aborted re-infection, then the boost of their antibody levels hints to a responsive adaptive immune system with the ability to control the disease.

While regular re-exposure may sustain protective antibodies, this is based on the predicament that new resistant viral variants will not arise. Although new variants are circulating already world-wide, the impact of the Spike/RBD mutations has still to be defined.^26–28^ Of note, our LV pseudo-particles bear the Wuhan-01 Spike while at the time of our study, the Italian epidemics was characterized by the predominance, if not exclusiveness, of the SARS-CoV-2 carrying the Spike D614G-variant.^29^ Nonetheless, our neutralization assay detected high titer nAbs in the majority of patients. This is a partially reassuring evidence, boding well for the vaccination campaign just started in Italy and worldwide with immunogens based on the original Chinese variant. However, should novel strains with nAbs escape mutation(s) arise, a plan of intervention would require updated Spike sequences for vaccination. Ongoing vaccination and re-infection studies combined with antibody kinetics to the Spike in natural infection will contribute to define correlates of protection and clarify factors that impact on adaptive immune-responses, which in turn would be beneficial for modelling the future COVID-19 dynamics.^30^

## Supporting information

Dispinseri_supplementary materials

## Data Availability

The datasets generated during and/or analysed during the current study are available from the corresponding author on reasonable request.

## Contributors

GS, VL, AC conceived and designed the study.

GS, SD, VL wrote the first draft with contributions from AC, DN, LP, MB.

VL, LP, GS did the statistical analysis.

SD, MT, FS, MS, MFP, MB, MLdA, ACan, GV, CB, EB, IM did the laboratory tests.

SD, MB, VL, GS, LP, CT did the data organization, data storage and quality control.

CT, SD, MS, MT oversaw sample handling and storage.

FC, GS oversaw institutional review board submissions, approval, and study oversight.

GS, LP, VL, FC, AC, DN acquired funding.

All authors reviewed the manuscript for intellectual content and assisted in data interpretation.

VL, GS, DN, AC, SD prepared the manuscript for submission.

All authors confirm that they had full access to all the data in the study and accept responsibility to submit for publication.

## Acknowledgements

The work performed at IRCCS Ospedale San Raffaele (OSR) was funded by Program Project COVID-19 OSR-UniSR and Ministero della Salute (COVID-2020-12371617). The work by Viral Evolution and Transmission Unit, OSR, and Istituto Superiore di Sanità (ISS) was funded by EAVI2020, the European Union’s Horizon 2020 research and innovation programme under grant agreement No. 681137. ISS received support in part by NATO multi-year Project No. G5817 “New and Validated Tools for the Diagnosis and follow-up of SARS-CoV-2 Infected Individuals” and by ISS internal funds. We thank the Fondation Dormeur, Vaduz for the donation to Viral Evolution and Transmission Unit for laboratory instruments relevant to this project. The following reagent was obtained through the NIH AIDS Reagent Program, Division of AIDS, NIAID, NIH: Anti-HIV-1 SF2 p24 Polyclonal. We thank Florian Krammer, Mount Sinai, for kindly donating SARS-CoV-2 Spike plasmids.

Special acknowledement goes to the COVID-BioB team^#^ and healthcare workers at OSR that made this work possible.

## Data Availability Statement

The raw data supporting the conclusions of this article will be made available by the authors, without undue reservation, with publication.

## ^#^COVID-BioB study team and collaborators

Affiliation is at IRCCS Ospedale San Raffaele, Milan, Italy.

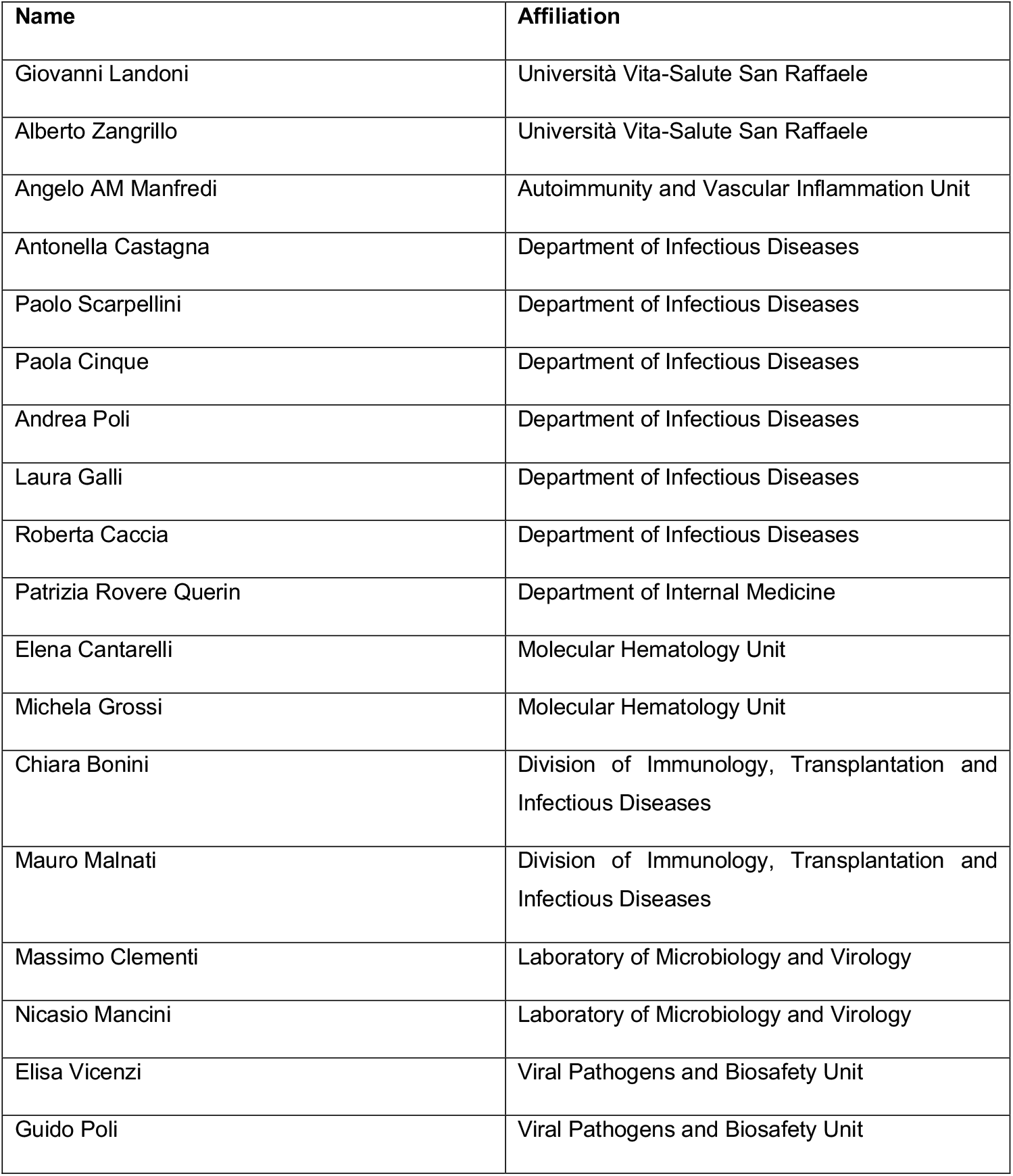

## Bibliography

1 Wu Z, McGoogan JM. Characteristics of and Important Lessons From the Coronavirus Disease 2019 (COVID-19) Outbreak in China: Summary of a Report of 72?314 Cases From the Chinese Center for Disease Control and Prevention. JAMA 2020; 323: 1239–42.

2 Long Q-X, Liu B-Z, Deng H-J, et al. Antibody responses to SARS-CoV-2 in patients with COVID-19. Nat Med 2020; 26: 845–8.

3 Zohar T, Loos C, Fischinger S, et al. Compromised Humoral Functional Evolution Tracks with SARS-CoV-2 Mortality. Cell 2020; 183: 1508-1519.e12.

4 Secchi M, Bazzigaluppi E, Brigatti C, et al. COVID-19 survival associates with the immunoglobulin response to the SARS-CoV-2 spike Receptor Binding Domain. J Clin Invest 2020; published online Sept 29. DOI:10.1172/jci142804.

5 Weinreich DM, Sivapalasingam S, Norton T, et al. REGN-COV2, a Neutralizing Antibody Cocktail, in Outpatients with Covid-19. N Engl J Med 2020; published online Dec 17. DOI:10.1056/NEJMoa2035002.

6 Libster R, Pérez Marc G, Wappner D, et al. Early High-Titer Plasma Therapy to Prevent Severe Covid-19 in Older Adults. N Engl J Med 2021; published online Jan 6. DOI:10.1056/NEJMoa2033700.

7 Chen P, Nirula A, Heller B, et al. SARS-CoV-2 Neutralizing Antibody LY-CoV555 in Outpatients with Covid-19. N Engl J Med 2020; published online Oct 28. DOI:10.1056/nejmoa2029849.

8 Wu F, Liu M, Wang A, et al. Evaluating the Association of Clinical Characteristics With Neutralizing Antibody Levels in Patients Who Have Recovered From Mild COVID-19 in Shanghai, China. JAMA Intern Med 2020; 180: 1356–62.

9 Wang X, Guo X, Xin Q, et al. Neutralizing Antibody Responses to Severe Acute Respiratory Syndrome Coronavirus 2 in Coronavirus Disease 2019 Inpatients and Convalescent Patients. Clin Infect Dis 2020; published online June 4. DOI:10.1093/cid/ciaa721.

10 Temperton NJ, Chan PK, Simmons G, et al. Longitudinally profiling neutralizing antibody response to SARS coronavirus with pseudotypes. Emerg Infect Dis 2005; 11: 411–6.

11 Choe PG, RAPM Perera, Park WB, et al. MERS-CoV antibody responses 1 year after symptom onset, South Korea, 2015. Emerg Infect Dis 2017; 23: 1079–84.

12 Wang K, Long Q-X, Deng H-J, et al. Longitudinal Dynamics of the Neutralizing Antibody Response to Severe Acute Respiratory Syndrome Coronavirus 2 (SARS-CoV-2) Infection. Clin Infect Dis 2020; published online Aug 3. DOI:10.1093/cid/ciaa1143.

13 Seow J, Graham C, Merrick B, et al. Longitudinal observation and decline of neutralizing antibody responses in the three months following SARS-CoV-2 infection in humans. Nat Microbiol 2020. DOI:10.1038/s41564-020-00813-8.

14 Dan JM, Mateus J, Kato Y, et al. Immunological memory to SARS-CoV-2 assessed for up to 8 months Mafter infection. Science (80-) 2021;: eabf4063.

15 Rovere-Querini P, Tresoldi C, Conte C, et al. Biobanking for COVID-19 research. Panminerva Med 2020; published online Oct 19. DOI:10.23736/S0031-0808.20.04168-3.

16 Fenyö EM, Heath A, Dispinseri S, et al. International network for comparison of HIV neutralization assays: The NeutNet report. PLoS One 2009; 4. DOI:10.1371/journal.pone.0004505.

17 Team RC. R: A Language and Environment for Statistical Computing. https://www.r-project.org.

18 Giroglou T, Cinatl Jr J, Rabenau H, et al. Retroviral vectors pseudotyped with severe acute respiratory syndrome coronavirus S protein. J Virol 2004; 78: 9007–15.

19 Sadasivan J, Singh M, JDAS Sarma. Cytoplasmic tail of coronavirus spike protein has intracellular targeting signals. J Biosci 2017; 42: 231–44.

20 Wang B-Z, Liu W, Kang S-M, et al. Incorporation of High Levels of Chimeric Human Immunodeficiency Virus Envelope Glycoproteins into Virus-Like Particles. J Virol 2007; 81: 10869–78.

21 Anderson EM, Goodwin EC, Verma A, et al. Seasonal human coronavirus antibodies are boosted upon SARS-CoV-2 infection but not associated with protection. medRxiv Prepr Serv Heal Sci 2020; published online Nov 10. DOI:10.1101/2020.11.06.20227215.

22 Aydillo T, Rombauts A, Stadlbauer D, et al. Antibody Immunological Imprinting on COVID-19 Patients. medRxiv 2020;: 2020.10.14.20212662.

23 FDA. Coronavirus (COVID-19) Update: FDA Authorizes Monoclonal Antibodies for Treatment of COVID-19. https://www.fda.gov/news-events/press-announcements/coronavirus-covid-19-update-fda-authorizes-monoclonal-antibodies-treatment-covid-19.

24 Gaebler C, Wang Z, Lorenzi JCC, et al. Evolution of Antibody Immunity to SARS-CoV-2. BioRxiv 2021;: 2020.11.03.367391.

25 Ministero della salute. Monitoraggio settimanale Covid-19, report 21-27 settembre. http://www.salute.gov.it/portale/nuovocoronavirus/dettaglioNotizieNuovoCoronavirus.jsp?lingua=italiano&menu=notizie&p=dalministero&id=5093..

26 Weissman D, Alameh MG, de Silva T, et al. D614G Spike Mutation Increases SARS CoV-2 Susceptibility to Neutralization. Cell Host Microbe 2020. DOI:10.1016/j.chom.2020.11.012.

27 Xie X, Zou J, Fontes-Garfias CR, et al. Neutralization of N501Y mutant SARS-CoV-2 by BNT162b2 vaccine-elicited sera. bioRxiv 2021;: 2021.01.07.425740.

28 Tegally H, Wilkinson E, Giovanetti M, et al. Emergence and rapid spread of a new severe acute respiratory syndrome-related coronavirus 2 (SARS-CoV-2) lineage with multiple spike mutations in South Africa. medRxiv 2020;: 2020.12.21.20248640.

29 Korber B, Fischer WM, Gnanakaran S, et al. Tracking Changes in SARS-CoV-2 Spike: Evidence that D614G Increases Infectivity of the COVID-19 Virus. Cell 2020; 182: 812-827.e19.

30 Saad-Roy CM, Wagner CE, Baker RE, et al. Immune life history, vaccination, and the dynamics of SARS-CoV-2 over the next 5 years. Science (80-) 2020; 370: 811.

