## Supplementary material for "COVID-BioB Cohort Study: the neutralizing antibody response to SARS-CoV-2 in symptomatic COVID-19 is persistent and critical for virus control and survival": Dispinseri_supplementary materials

by Stefania Dispinseri et al.

This file includes:

- Material and methods
- Results
- Tables S1-4
- Figures S1-8

### MATERIALS AND METHODS

*Study population and Data sources*

The study participants are part of the COVID-19 clinical-biological cohort study (COVID-BioB, ClinicalTrialsgov identifier NCT04318366) at the IRCCS San Raffaele Hospital, reviewed and approved by the Review Board (RB) (protocol number 34/int/2020). Patients admitted to the Emergency department (ED) from February 25th, 2020 were enrolled according to the protocol described in.[1] All enrolled participants gave written, informed consent.

A confirmed infection case was defined as a SARS-CoV-2 positive real-time reverse-transcriptase polymerase chain reaction (RT-PCR) from a nasal-pharyngeal swab and/or symptoms and radiological findings suggestive of COVID-19 pneumonia. Routine blood tests included complete blood count with differential, renal and liver function tests, C-reactive protein (CRP) and lactate dehydrogenase (LDH), while serum ferritin, D-Dimer and interleukin-(IL-6) were on request for clinical reasons (table S1). The clinical and laboratory data was collected from medical chart review or directly by patient interview, crosschecked for accuracy by data managers and clinicians and entered in a dedicated electronic case record form (eCRF) developed on site for the COVID-BioB study.

Biological material for COVID-BioB included a dedicated serum sample collected during attendance at the hospital, which took place at the ED or at the ward (called the in-hospital sample). Follow-up serum samples were collected at the post-COVID19 out-patient clinic during the one month, 3 and 6 months or subsequent visits after discharge. All serum samples were coded and anonymously processed until disclosure of the COVID-19 status for the immunology studies concluded by November 25th 2020. The immunological study “Role of the immune response in the infection with SARS-COV-2 and in the pathogenesis of COVID-19”, ImmCOVID, received the approval from the RB (protocol number 68/INT/2020). A total of 362 sera of COVID-19 confirmed patients were tested for SARS-CoV-2 neutralizing antibody (nAbs) response and antibody binding to SARS-CoV-2, HCoV-OC43, HCoV-HKU1 and H1N1 Influenza. Another 17 individuals admitted to the hospital during the same period with respiratory symptoms, but no SARS-CoV-2 diagnosis, were included.

*Plasmids*

Schematic representation of plasmids used in this report is shown in figure S1. The SIV-based self-inactivating (SIN) lentiviral transfer vector expressing luciferase (Luc) reporte gene (pGAE-LucW), the SIV-based packaging plasmid (pAdSIV3+) producing the proteins necessary for particles formation and the phCMV-VSV.G plasmid producing the pseudotyping vesicular stomatitis virus envelope glycoprotein G (VSV.G) have been already described.[2] For construction of pSpike plasmid, the full length codon optimized SARS-CoV-2 Spike protein open reading frame (ORF) (GenBank: NC_045512.2) was chemically synthesized, inserted into pUC57 plasmid (TwinHelix, Milan, Italy), excised with AgeI/SalI restriction enzymes and cloned into pEGFP-C3 plasmid (Clontech Mountain View, CA, USA). For construction of pSpike-C3 plasmid expressing the wild-type Spike ORF with a 21bp deletion at the cytoplasmic tail, the pSpike plasmid was digested with BamHI restriction enzyme and self-ligated.

*Production of lentiviral vector pseudotypes expressing luciferase*

Lenti-X human embryonic kidney 293T cell line (Clontech) was maintained in Dulbecco's Modified Eagles (DMEM) High glucose 4.5 g/L (Gibco, Life Technologies Italia, Monza, Italy) supplemented with 10% fetal calf serum (Corning, Mediatech inc. Manassas, VA, USA) and 100 units/mL penicillin/streptomycin (Gibco). For production of Lentiviral vectors expressing luciferase (LV-Luc), 3.5 x $10^6$ 293T Lenti-X cells were seeded on 10 cm Petri dishes (Corning Incorporated - Life Sciences, Oneonta, NY, USA) and transiently transfected with plasmids pGAE-LucW, pADSIV3+ and pseudotyping plasmid (phCMV-VSV.G, pSpike or pSpike-C3) using the JetPrime transfection kit (Polyplus Transfection Illkirch, France) following the manufacture's recommendations using a 1:2:1 ratio (transfer vector : packaging plasmid : envelope/Spike plasmid). At 48 h from transfections, culture supernatants containing the LV-Luc pseudotypes (LV-Luc/Spike-C3, LV-Luc/Spike and LV-Luc/VSV.G) were collected, cleared from cellular debris by low-speed centrifugation, passed through a 0.45 μm pore size filter unit (Millipore, Billerica, MA) and stored in 1 mL aliquotes at -80°C until use. To produce Spike-pseudotyped LV-Luc stocks for Western blot analysis, vector containing supernatants were concentrated by ultracentrifugation (Beckman Coulter, Fullerton, CA, USA) on a 20% sucrose cushion (Sigma Chemical Co., St. Louis, MO, USA) at 23.000 rpm for 2.5 h at 4°C using a SW28 swinging bucket rotor (Beckman). Purified pelleted vector particles were resuspended in 1× phosphate-buffered saline (PBS, Gibco, Life Technologies Italia, Monza, Italy) and stored at −80°C until use. Each LV stock was measured by the reverse transcriptase (RT) activity assay[3] and titrated for luciferase activity on ACE2+ cells, as described below.

*Flow cytometry to evaluate Spike expression*

293T Lenti-X cells were seeded in 10 cm plates and transfected with 5 μg of pSpike or pSpike-C3 plasmid using the JetPrime transfection kit (Polyplus Transfection). At 48 h after transfection, cells were collected, washed with PBS1X (Gibco) and fixed with 1% paraformaldehyde. Spike expression was evaluated by flow cytometry with anti-Spike S2 rabbit polyclonal Ab (Sinobiological, Sino Biological Europe GmbH, Germany) and PE-conjugated donkey anti-rabbit IgG (Biolegend, San Diego, CA, USA) using the FACSCalibur (BD Biosciences, Milan, Italy), and data were analyzed by CellQuest Pro software (BD).

*Western Blot (WB)*

To evaluate Spike presence on LV particles, pellets of concentrated preparations were resuspended and lysed in SDS loading buffer. Lysed virions were separated on 10% SDS polyacrylamide gel under reducing conditions and transferred to a nitrocellulose membrane with a Trans-Blot Turbo System (Bio-Rad, CA, USA). Filters were saturated for 1 h with 5% nonfat dry milk in TSBS (TBS with 0.1% Tween 20) and then incubated with polyclonal anti-Spike (Sino Biological, USA, Cat. 40590-T62) or anti-Gag antibody (NIH Repository Reagents, Cat. #4250), as primary antibodies, for 2 hs at room temperature, followed by incubation for 1 h at room temperature with an anti-rabbit horse radish peroxidase (HRP)-conjugated IgG (Bio-Rad). The immunocomplexes were visualized using chemiluminescence ECL detection system (WesternBright ECL, Advansta).

*Cell lines*

Caco2 (human, colon), Calu3 (human, lung), HUH-7 (human, liver), VEROE6 (African green monkeys, kidney), and 293T (human, kidney) cell lines were grown in Dulbecco's Modified Eagle Medium (DMEM) with Ultraglutamine (EuroClone, Italy) supplemented with 10% (or 20% for Calu3) Fetal Bovine Serum (FBS, EuroClone, Italy), 10 mM of HEPES (1M) (EuroClone, Italy), 1% Penicillin-Streptomycin (100X) (EuroClone, Italy), 1% Non Essential Aminoacids (NEAA) (100X) (EuroClone, Italy), 1% NA Pyruvate (100mM) (Euroclone, Italy); Raji (human B cell lymphoma) cells were grown in RPMI 1640 medium with ultraglutamine (EuroClone, Italy) supplemented with 10% FBS, 10 mM of HEPES (1M), 1% Penicillin-Streptomycin (100X), 1% NEAA (100X), 1% NA Pyruvate (100 mM). All cell lines were cultured at 37ºC 5% $CO_2$ and passaged every 2-3 days.

*Flow cytometry to evaluate ACE2 expression*

To verify the expression of ACE2 on the cell lines, 200.000 of each cell type were stained with mouse anti-human ACE2 (Millipore, cod. MAB5676) for 30 min at +4ºC followed by Phycoerythrin conjugated goat anti-mouse secondary antibody (SouthernBiotech, USA) for 30 min at +4ºC. The samples were acquired at Navios flow cytometer (Beckman Coulter, Inc.) and analyzed using FlowJo version 8.8.7 (Tree Star).

*Viral cell infectivity assay*

Infection of a panel of cell lines was performed with LV-Luc/Spike and LV-Luc/Spike-C3 in replicates of three serial initial 2-fold dilution. Two or more cell densities were used for each cell line. Caco2 (40.000 up to 250.000 cells), Calu3 (50.000 up to 250.000 cells), HUH7 (7.000 or 15.000 cells), VEROE6 (20.000, 50.000 or 100.000 cells), and Raji (70.000, 100.000 or 200.000 cells) were seeded at the indicate number per well in growth medium in 96-well culture plates and incubated at 37ºC 5% $CO_2$ until cells were well-settled. Culture fluid was removed and 100 µL of each viral dilution added to the cells. After 24 and 48h of incubation at 37ºC/5% $CO_2$, the activity of firefly luciferase was measured with a Luc reporter gene assay system reagent (Bright-Glo, Promega, Madison, Wisconsin, US): 100 µL of Bright-Glo was added to each well and after 2 min incubation at room temperature in the dark to allow cell lysis, thereafter 100 µL of cell lysate was transferred to 96-well white plate (Perkin-Elmer, Life Sciences, Shelton, CT) for measurements of relative light units (RLU) of luminescence using a Mithras luminometer (Berthold, Germany).

*Viral titration assay*

Titration of the LVs was performed in VEROE6 cell cultures. LV-Luc/Spike-C3 was titred performing five replicates of serial 2-fold initial dilutions, and control LV-Luc/VSV.G was titred performing 10-fold initial dilution in duplicate in growth medium in 96-well culture plates. Freshly trypsinized VEROE6 cells were added to each well in growth medium and incubated at 37ºC until cells were well-settled. Culture supernatant was removed and replaced with 100 µL of each virus dilution and further cultured. After 48 h 100 µL of Bright-Glo was added to each well and incubated for 2 min at room temperature in the dark to allow cell lysis; 100 µL of cell lysate was transferred to 96-well white plate and RLU measured using a Mithras luminometer. The dilution of the virus supernatant providing 150.000-200.000 RLU was used in the neutralization assay.

*LV based neutralization assay*

Pseudoviruses LV-Luc/Spike-C3 and control LV-Luc/VSV.G were used in the neutralization assay. Serum was heat inactivated by incubation at 56°C for 30 min. LV-Luc/Spike-C3 supernatant was incubated with four or six 3-fold serial dilutions starting with 1/40 of each heat inactivate serum, for 30 min at 37°C in 96-well plates. VEROE6 at a density of 20.000 cells/well were added to flat bottom 96-well plates in growth medium. After 4 h of incubation at 37°C to allow cells to well-settle to the plate, culture supernatant was discarded, and the virus-serum mixture added in duplicates to the cells. Plates were incubated at 37°C for 48 h, and thereafter the procedure followed the same steps as for the viral titration assay described above, until the measurement of luminescence in a Mithras instrument.

Virus controls were added to each 96-well test plate and included 5 wells with LV-Luc/Spike-C3 and cells. Additional controls were four 3-fold serial dilutions of two sera with a known SARS-CoV-2 neutralization titer and of a negative serum added in duplicate to each plate. The 50% and 75% inhibitory serum dilution (ID50 and ID75) were calculated with a linear interpolation method using the mean of the duplicate responses.[4] Neutralization was expressed as the reciprocal of the serum dilution giving 50% inhibition of RLU compared to the mean of the virus control wells. An ID50 below 1/40 serum dilution was considered negative and a value of 10 ascribed for statistical analysis.

A test was valid if the percent coefficient of variation (%CV) RLUs of the virus control wells was ≤30%, and the value of both positive control sera was within a 2-fold range of the median inter-assay ID50 value. A test sample result was discarded and the serum re-tested when the percent difference between duplicate wells was >30% for sample dilutions that yielded at least 40% inhibition.

To exclude any unspecific inhibition each serum was tested in duplicate at 1/40 dilution with LV-Luc/VSV.G in the neutralization assay, as described above. Serum with >50% VSV inhibition compared to virus control was discarded from any further analysis.

*Fluid-phase luciferase immune precipitation (LIPS) assay*

The assay was previously standardized and published in [5]. We produced several recombinant monomeric or multimeric SARS-CoV-2 proteins tagged with a Nanoluciferase reporter (Promega, Madison, Wisconsin, USA): the whole spike glycoprotein S1+S2 (a derivative of the previously described trimeric S1+S2 spike protein[6] or its parts the S2 protein and the glycoprotein RBD, the nucleocapsid protein (NP), the spike S2 proteins of the HCoV-OC43 and HCoV-HKU1 betacoronaviruses and the hemagglutinin HA1 protein of the 2009 H1N1 pandemic flu virus (figure S2). Recombinant nanoluciferase-tagged antigens were expressed by transient transfection into Expi293F™ cells (Expi293™ Expression System, Thermo Fisher Scientific Life Technologies, Carlsbad, CA, USA) according to the manufacturer's instructions. Upon harvesting, recombinant proteins were aliquoted and stored frozen at -80°C.

For antibody measurement by LIPS, the antigen of interest was thawed, diluted in 20 mM Tris Buffer, 150 mM NaCl, 0.5% Tween-20, pH 7.4 (TBST) buffer and adjusted to achieve a luciferase activity corresponding to a final concentration of 4 x $10^6$ Light Units (LU)/25 µL. Serum was then seeded (5 µL for IgM, 1 µL for IgG or IgA measurements, respectively) was seeded into the well of a 96-deep-well plate (Beckman Coulter Inc., Brea, CA, USA) added with 25 μL of the diluted antigen preparation, and incubated for 2 h at room temperature. For IgG antibody measurement, immunocomplexes were then captured with 5 μL of a 50% weight/volume blocked rProtein A slurry (GE Healthcare Europe GmbH, Freiburg, Germany) for 1 h at 4°C with shaking. Plates were washed 5 times by sequentially dispensing 750 μL/well of TBST, followed by centrifugation at 500g for 3 min at 4°C and removal of the wash buffer using a micro-plate washer/dispenser (BioTek Instruments Inc., Winooski, VT, USA). For IgM or IgA antibody measurements, rProtein A was replaced with 5 μL of goat anti-human IgM- or anti-human IgA agarose (Merck Life Sciences, Milano, Italy), respectively.

After washing, the resin pellets were transferred to an OptiPlate™ 96-well plate (PerkinElmer, Waltham, MA, USA), added with 40 μL/well of Nano-Glo® substrate (Promega) and the recovered luciferase activity was measured over 2 sec/well in a Berthold Centro XS3 luminometer (Berthold Technologies GmbH & Co. KG, Bad Wildbad, Germany). Raw data were converted to Arbitrary Units (AU) using either a local positive serum as index or serial dilution of a SARS-COV-2 Spike protein antibody positive serum (a kind gift of Prof. Ezio Bonifacio, Dresden, Germany). For antibody titrations, the sera that bound recombinant antigens above the linear range of the assay were serially diluted (1/10, 1/100, 1/1000) in TBST, re-tested until binding fell into the linear range and the calculated AU corrected by multiplying for the corresponding dilution factor.

Thresholds for antibody positivity were established upon a QQ plot analysis by selecting AU values at which the distribution of calculated AU deviated from normality. For ubiquitously present antibody responses like those against the 2009 pandemic flu HA and the HCoV-OC43 and HCoV-HKU1 S2 spike proteins, subjects were binned into terciles as follows: OC43 first tercile 0-270.3 AU, second tercile 270.4 – 1469.7 AU, third tercile > 1469.7 AU; HKU1 first tercile 0-312.3 AU, second tercile 312.4 – 1471.9 AU, and third tercile > 1471.9 AU.

### RESULTS

#### Viral Infectivity

LVs were used to infect a panel of four epithelial cell lines of human or macaque intestinal or pulmonary origin, which express ACE2, a receptor involved in SARS-CoV-2 cell entry, as well as one B cell line. Two or more cell densities for each cell line, as indicated in the methods section, were seeded in duplicates and cultured to test vitality and density after 24 and 48 h. 20.000, 50.000 or 100.000 VEROE6, 40.000 to 250.000 Caco2, 7.000 or 15.000 HUH7, 50.000 to 250.000 Calu3 and 70.000, 100.000 or 200.000 Raji cells were seeded in growth medium. Trypan blue (Sigma) vitality count showed that 90%-100% of cells were alive at all densities seeded. Evaluation of cell growth at optical microscopy demonstrated that confluence was reached after 24 h when 50.000 VEROE6, 70.000 Caco2, 100.000 Calu3 or 200.000 Raji were seeded and after 48 h, when 20.000 VEROE6, 70.000 Caco2, 50.000 Calu3 and 70.000 Raji were seeded. HUH7 achieved the expected confluence at 24 and 48 h when 7.000 cells were seeded. Thus, 20.000 VEROE6, 70.000 Caco2, 50.000 Calu3, 7.000 HUH7 and 70.000 Raji cells were used for further infectivity experiments to be evaluated for luminescence at 48 h.

The cell lines were further tested for infectivity with the LVs, normalized for RT-activity, at increasing dilutions. LV-Luc/Spike was always, at all dilutions tested, less infective than LV-Luc/Spike-C3. However, the difference was more pronounced in VEROE6 cells (50-fold), followed by HUH7 cells (9-fold) and Caco2 (3-fold), while in Calu3 cells only Luc/Spike-C3 showed some low level infection (maximum 250 RLU). As expected Raji cells were not infected. The assay background level did not exceed 100 RLU for all the cell lines tested (figure 1C).

A titration was performed in VEROE6 cells with five replicates of 2-fold dilutions starting with 1/4 of LV-Luc/Spike-C3 to determine the input virus dilution for the neutralization assay. Two different LV-Luc/Spike-C3 productions were compared and gave similar results (data not shown). The virus dilution with 150.000-200.000 RLU, which had a CV below 30%, was selected for the neutralization assay.

Serum samples from SARC-CoV-2 infected and uninfected individuals (collected pre-2019 and during the epidemic) were used to set the neutralization assay. Sera from 19 uninfected individuals, tested at 1/20 and 1/40 dilution, gave results in the range of the background RLU levels. Conscious that some very low level (between 1/20 and 1/39) neutralization titers would be missed, we chose the 1/40 dilution as the first suitable serum dilution for the assay, with the intention to save valuable biological specimens. None of the sera of infected and uninfected individuals, which had detectable IgG to HCoVs OC43 and HKU1 and absent IgG to RBD, showed an inhibition in the neutralization assay, and thus, excluded an unspecific inhibition of SARS-CoV-2 by these antibodies. The COVID-19 sera were repeatedly tested to set the intra- and inter-assay variation and thus, define the validity criteria, as described in Material and Methods.

*Antibody response of not hospitalized COVID-19 patients*

As previously reported, antibody persistence may vary according to the degree of the disease severity.[7] In our study all patients were symptomatic at disease onset, though 26 did not require hospitalization (table S4). The latter characteristics were different from the overall cohort, with the majority being younger females (61.5%, median age 49.5 years) without co-morbidities (80.8%) although with symptoms at disease onset similar to hospitalized patients, save for the absence of dyspnea. In non-hospitalized patients, the profile of nAbs and binding antibodies to the Spike protein was similar to that in hospitalize patients (figure S8). None of the 12 non-hospitalized patients tested during follow-up lost the nAb response until the 2$^{nd}$ and 3$^{rd}$ outpatient visit (range 76-218 days after sympotns onset).

### Bibliography


1 Rovere-Querini P, Tresoldi C, Conte C, *et al.* Biobanking for COVID-19 research. *Panminerva Med* 2020; published online Oct 19. DOI:10.23736/S0031-0808.20.04168-3.

2 Michelini Z, Galluzzo CM, Negri DRM, *et al.* Evaluation of HIV-1 integrase inhibitors on human primary macrophages using a luciferase-based single-cycle phenotypic assay. *J Virol Methods* 2010; **168**: 272–6.

3 Weiss S, König B, Morikawa Y, Jones I. Recombinant HIV-1 nucleocapsid protein p15 produced

as a fusion protein with glutathione S-transferase in Escherichia coli mediates dimerization and enhances reverse transcription of retroviral RNA. *Gene* 1992; **121**: 203–12.

4 Fenyö EM, Heath A, Dispinseri S, *et al.* International network for comparison of HIV neutralization assays: The NeutNet report. *PLoS One* 2009; **4**. DOI:10.1371/journal.pone.0004505.

5 Secchi M, Bazzigaluppi E, Brigatti C, *et al.* COVID-19 survival associates with the immunoglobulin response to the SARS-CoV-2 spike Receptor Binding Domain. *J Clin Invest* 2020; published online Sept 29. DOI:10.1172/jci142804.

6 Amanat F, Stadlbauer D, Strohmeier S, *et al.* A serological assay to detect SARS-CoV-2 seroconversion in humans. *Nat Med* 2020; **26**: 1033–6.

7 Wajnberg A, Amanat F, Firpo A, *et al.* Robust neutralizing antibodies to SARS-CoV-2 infection persist for months. *Science (80- )* 2020; : eabd7728.

**Table S1: Laboratory values at first Biobank blood sampling of the COVID-19 study population.**

| Characteristics | COVID-19 patients | Missing data |
|---|---|---|
| N | 162 | |
| Median time (days) from symptom onset to first sampling for Biobank | 11.5 (7-18) | 0 |
| Laboratory values at time of first blood sampling: | Median (IQR) | |
| - White Blood Cells, x$10^9$/L | 6.9 (5.2-9.8) | 17 |
| - Lymphocyte (Ly) count, x$10^9$/L | 1.1 (0.7-1.5) | 25 |
| - Neutrophil (N) count, x$10^9$/L | 4.8 (3.5-7.5) | 25 |
| - Monocyte count, x$10^9$/L | 0.5 (0.4-0.7) | 25 |
| - N/Ly ratio | 4.67 (2.55-8.75) | 25 |
| - Haemoglobin, g/dL | 13.1 (11.6-14.6) | 17 |
| - Platelet count, x$10^9$/L | 236 (182-323) | 17 |
| - Bilirubin total, mg/dL | 0.56 (0.37-0.86) | 45 |
| - ALT, U/L | 41 (23.5-66.5) | 35 |
| - AST, U/L | 44 (31-60) | 34 |
| - Creatinine, mg/dL | 0.97 (0.77-1.34) | 23 |
| - LDH, U/L | 355 (273-428) | 39 |
| - CRP, mg/L | 54.6 (18.75-118.1) | 19 |
| - D-Dimer, µg/mL | 1.11 (0.53-2.45) | 104 |
| - IL-6, pg/mL | 33 (16.7-81) | 117 |
| - Ferritin, ng/mL | 1222 (597-1701) | 103 |

**Footnote to Table S1:** ALT: Alanine Amino Transferase; AST: Aspartate Amino Transaminase; LDH: Lactate dehydrogenase; CRP: C-reactive protein.

**TABLE S2: SARS-CoV-2 Antibody responses according to week from symptoms onset**

| | Weeks after symptoms onset | | | | | |
|---|---|---|---|---|---|---|
| | 1-2 | 3-4 | 5-8 | 9-16 | 17-36 | Overall |
| **Samples (N=)** | 101 | 43 | 76 | 76 | 66 | 362 |
| **SARS-CoV-2 Neutralization** | | | | | | |
| Mean (SD) | 3500 (8600) | 45300 (252000) | 9950 (24400) | 2160 (3910) | 1460 (1910) | 9190 (87900) |
| Median | 238 | 2640 | 3500 | 869 | 660 | 1080 |
| [Min, Max] | [10, 61500] | [10, 1660000] | [10, 161000] | [10, 28000] | [10, 10900] | [10, 1660000] |
| **IgG_RBD** (AU) | | | | | | |
| Mean (SD) | 13.0 (29.4) | 165 (296) | 2670 (5710) | 4360 (7570) | 6100 (6640) | 2610 (5660) |
| Median | 1.40 | 43.0 | 1060 | 2010 | 3830 | 333 |
| [Min, Max] | [0.00296, 163] | [0.00859, 1130] | [0.116, 46500] | [0.0453, 36800] | [0.0171, 31500] | [0.00296, 46500] |
| **IgG_ S1S2** (AU) | | | | | | |
| Mean (SD) | 37.5 (50.6) | 1370 (4040) | 8850 (8960) | 8080 (8660) | 7310 (6970) | 5060 (7580) |
| Median | 8.66 | 82.3 | 6420 | 5350 | 6650 | 1080 |
| [Min, Max] | [0.0228, 258] | [0.379, 23400] | [0.160, 46900] | [6.65, 33400] | [0.112, 33400] | [0.0228, 46900] |
| **IgG_ S2** (AU) | | | | | | |
| Mean (SD) | 349 (1760) | 818 (3030) | 1550 (4050) | 1640 (1670) | 3550 (3930) | 1510 (3140) |
| Median | 7.81 | 159 | 429 | 1090 | 2740 | 312 |
| [Min, Max] | [0.00373, 16000] | [0.00885, 18800] | [0.0289, 31000] | [0.0159, 6400] | [0.0169, 25600] | [0.00373, 31000] |
| **IgG_ NP** (AU) | | | | | | |
| Mean (SD) | 20.3 (19.7) | 31.0 (19.8) | 49.3 (18.9) | NA (NA) | NA (NA) | 28.3 (22.4) |
| Median | 12.4 | 35.7 | 53.1 | | | 29.4 |
| [Min, Max] | [0.237, 65.9] | 0.313, 65.6] | [0.443, 75.4] | NA (NA) | NA (NA) | [0.237, 75.4] |
| missing | 1 (1%) | 4 (9.3%) | 43 (56.6%) | 100% | 100% | 190 (52.5%) |
| **Other Viruses** | | | | | | |
| **IgG_OC43 S2** (AU) | | | | | | |
| Mean (SD) | 1430 (2040) | 2310 (2370) | 3490 (2430) | 2610 (2070) | 1630 (1190) | 2290 (2220) |
| Median | 492 | 1220 | | 2350 | 1320 | 1440 |
| [Min, Max] | [10.2, 10200] | [0.307, 8410] | 3410 | [29.8, 8340] | [57.6, 4420] | [0.307, 10200] |

| | | | | | | |
|---|---|---|---|---|---|---|
| missing | 0% | 0% | [60.1, 8780]<br>0% | 0% | 18 (27.3%) | 18 (5.0%) |
| **IgG_HKU1 S2** (AU)<br>Mean (SD)<br>Median<br>[Min, Max]<br>missing | <br>1760 (2550)<br>587<br>[21.4, 13500]<br>0% | <br>3600 (3030)<br>3410<br>[92.8, 10500]<br>0% | <br>4650 (4060)<br>3240<br>[56.8, 15100]<br>0% | <br>3580 (3020)<br>2910<br>[2.73, 12800]<br>0% | <br>2120 (1780)<br>1590<br>[108, 7310]<br>18 (27.3%) | <br>3080 (3210)<br>1930<br>[2.73, 15100]<br>18 (5.0%) |
| **IgG_FLU HA** (AU)<br>Mean (SD)<br>Median<br>[Min, Max]<br>missing | <br>18600 (25600)<br>7040<br>[72.6, 108000]<br>1 (1%) | <br>12500 (20900)<br>4750<br>[172, 117000]<br>4 (9.3%) | <br>13300 (15100)<br>6940<br>[311, 58200]<br>43 (56.6%) | <br>NA (NA)<br><br>NA (NA)<br>(100%) | <br>NA (NA)<br><br>NA (NA)<br>100% | <br>16200 (23000)<br>6890<br>[72.6, 117000]<br>190 (52.5%) |
| **IgM_FLU HA** (AU)<br>Mean (SD)<br>Median<br>[Min, Max]<br>missing | <br>9.34 (15.4)<br>2.82<br>[0.328, 92.5]<br>1 (1%) | <br>11.0 (17.7)<br>3.52<br>[0.159, 90.8]<br>4 (9.3%) | <br>14.0 (23.3)<br>2.97<br>[0.377, 91.4]<br>44 (57.9%) | <br>NA (NA)<br><br>NA (NA)<br>(100%) | <br>NA (NA)<br><br>NA (NA)<br>100% | <br>10.6 (17.6)<br>3.12<br>[0.159, 92.5]<br>191 (52.8%) |
| **Days post symptoms onset**<br>Median [Min, Max] | 8.00 [1, 14] | 18.0 [15, 28] | 39.0 [30, 54] | 95.0 [57, 112] | 204 [114, 250] | 39.0 [1, 250] |
| **Age_years**<br>Median [Min, Max] | 63.0 [34, 94] | 63.0 [34, 88] | 61.5 [19, 87] | 59.5 [26, 87] | 61.0 [37, 87] | 62.0 [19, 94] |
| **Sex Male** | 67 (66.3%) | 31 (72.1%) | 48 (63.2%) | 49 (64.5%) | 41 (62.1%) | 236 (65.2%) |
| **Sample Category** | | | | | | |
| **In-hospital visit** | 101 (100%) | 39 (90.7%) | 10 (13.2%) | 0 (0%) | 0 (0%) | 150 (41.4%) |
| **1st follow-up visit** | 0 (0%) | 4 (9.3%) | 66 (86.8%) | 17 (22.4%) | 0 (0%) | 87 (24.0%) |
| **2nd follow-up visit** | 0 (0%) | 0 (0%) | 0 (0%) | 59 (77.6%) | 18 (27.3%) | 77 (21.3%) |
| **3rd follow-up visit** | 0 (0%) | 0 (0%) | 0 (0%) | 0 (0%) | 43 (65.2%) | 43 (11.9%) |

| | | | | | | |
|---|---|---|---|---|---|---|
| **4th follow-up visit** | 0 (0%) | 0 (0%) | 0 (0%) | 0 (0%) | 4 (6.1%) | 4 (1.1%) |
| **5th follow-up visit** | 0 (0%) | 0 (0%) | 0 (0%) | 0 (0%) | 1 (1.5%) | 1 (0.3%) |

**Footnote to Table S3:** Neutralization is expressed as the inverse of the serum dilution at which the ID50 was obtained in the LV-based neutralization assay. Absence of neutralization at the first dilution used (1/40) for the assay was ascribed a value of 10.

AU = arbitrary units.

**Table S3: Characteristics of COVID-19 patients grouped according to their SARS-CoV-2 neutralizing antibody response.**

| **SARS-CoV-2 neutralizing Ab score** | **Negative** | m | **Positive** | m | p Value |
|---|---|---|---|---|---|
| Number (N) | 43 | | 107 | | |
| Age, years | 74 (59-79) | 0 | 62 (51-70) | 0 | 0.003 |
| Comorbidities: | | | | | |
| - Hypertension | 26 (60.5%) | 0 | 45 (42.1%) | 0 | 0.048 |
| - CAD | 8 (18.6%) | | 13 (12.1%) | | 0.308 |
| - Diabetes | 10 (23.3%) | | 30 (28%) | | 0.684 |
| - COPD | 5 (11.6%) | | 1 (0.9%) | | 0.008 |
| - CKD | 15 (34.9%) | | 9 (8.4%) | | <0.001 |
| - Cancer | 10 (23.3%) | | 7 (6.5%) | | 0.008 |
| - Neurodegenerative disease | 3 (7%) | | 1 (1.9%) | | 0.142 |
| N of Comorbidities | | | | | |
| - None | 10 (23.3%) | 0 | 48 (44.9%) | 0 | 0.005 |
| - 1 | 6 (14%) | | 22 (20.6%) | | |
| - 2 | 14 (32.6%) | | 28 (26.2%) | | |
| - 3 | 9 (20.9%) | | 7 (6.5%) | | |
| - 4 | 4 (9.3%) | | 2 (1.9%) | | |
| Body Mass Index | 25.9 (23-29.7) | 9 | 28.3 (25.4-32.3) | 5 | 0.018 |
| <25 | 15 (44.1%) | | 19 (18.6%) | | 0.012 |
| 25-30 | 11 (32.4%) | | 45 (44.1%) | | |
| >30 | 8 (23.5%) | | 38 (37.3%) | | |
| Symptoms at disease onset: | | 1 | | 4 | |
| - general | | | | | |
| - fever | 71.4% | | 91.3% | | 0.004 |
| - headache | 21.4% | | 25.2% | | 0.541 |
| - fatigue/malaise | 28.6% | | 67% | | <0.001 |
| - myalgia/arthralgia | 26.2% | | 35.9% | | 0.331 |
| - respiratory | | | | | |
| - cough | 35.7% | | 71.8% | | <0.001 |
| - dyspnea | 47.6% | | 79.6% | | <0.001 |
| - sore throat | 11.9% | | 17.5% | | 0.464 |
| - chest pain | 14.3% | | 26.2% | | 0.133 |
| - gastrointestinal | | | | | |
| - diarrhea | 16.7% | | 31.1% | | 0.099 |
| - vomiting/nausea | 9.5% | | 14.6% | | 0.589 |
| - abdominal pain | 11.9% | | 7.8% | | 0.523 |
| - others | | | | | |
| - conjunctivitis | 4.8% | | 18.4% | | 0.038 |
| - hypo/anosmia | 16.7% | | 46.6% | | 0.001 |
| - hypo/dysgeusia | 21.5% | | 50.5% | | 0.002 |
| - skin rash | 7.1% | | 3.9% | | 0.413 |
| Median time (days) from symptoms onset: | | | | | |
| - to admission | 5 (3-12) | 0 | 10 (7-12) | 0 | <0.001 |
| - to blood sampling | 6 (4-12) | | 13 (9-16) | | <0.001 |
| Admitted to the hospital | 43/45 (95.5%) | 0 | 107/117 (91.4%) | 0 | <0.001 |
| - Discharged | 5/43 (11.6%) | | 11/107 (10.3%) | | |
| - Hospitalized | | | | | |
| - ≤7days | 6/43 (14%) | | 19/107 (17.8%) | | |
| - >7days | 16/43 (37.2%) | | 49/107 (45.8%) | | |
| - deceased | 15/43 (34.9%) | | 3/107 (2.8%) | | |
| - in need of ICU, recovered | 1/43 (2.3%) | | 14/107 (13.1%) | | |
| - in need of ICU, deceased | 0/43 (0%) | | 11/107 (7.3%) | | |
| Hospital stay, days (n=134) | 8 (4-24) | | 13 (6-21) | | 0.317 |
| Median time (days) from symptoms to negative RT-PCR swab (95%CI) | 46 (38-54) | 2 | 40 (37-43) | 5 | 0.041 |
| Median follow up, days (95%CI) | 194 (99-289) | 0 | 203 (198-208) | 0 | 0.521 |
| Laboratory values at time of first blood sampling: | | | | | |
| - White Blood Cells, x$10^9$/L | 6.2 (4.7-8.1) | 5 | 7.2 (5.5-10.3) | 12 | 0.028 |

| | | | | | |
|---|---|---|---|---|---|
| - Lymphocyte (Ly) count, x$10^9$/L | 0.95 (0.5-1.5) | 9 | 1.2 (0.8-1.5) | 16 | 0.088 |
| - Neutrophil (N) count, x$10^9$/L | 4.3 (2.65-6.75) | 9 | 4.9 (3.7-7.8) | 16 | 0.085 |
| - Monocyte count, x$10^9$/L | 0.4 (0.3-0.6) | 9 | 0.6 (0.4-0.8) | 16 | 0.009 |
| - N/Ly ratio | 4.2 (2.46-8.25) | 9 | 4.92 (2.55-9.04) | 16 | 0.986 |
| - Haemoglobin, g/dL | 11.95 (9.72-14.05) | 5 | 13.5 (12.1-14.8) | 12 | 0.001 |
| - Platelet count, x$10^9$/L | 187 (109-234) | 5 | 256 (199-355) | 12 | <0.001 |
| - Bilirubin total, mg/dL | 0.44 (0.36-0.72) | 13 | 0.65 (0.38-0.98) | 32 | 0.053 |
| - ALT, U/L | 30 (18.5-34.5) | 12 | 47 (26-75) | 23 | <0.001 |
| - AST, U/L | 36 (26-53) | 12 | 46 (33-64) | 22 | 0.045 |
| - Creatinine, mg/dL | 1.22 (0.87-1.81) | 7 | 0.95 (0.76-1.24) | 16 | 0.002 |
| - LDH, U/L | 282 (232-409) | 12 | 368.5 (294-441) | 27 | 0.007 |
| - CRP, mg/L | 46.4 (20.4-99) | 7 | 62.6 (18.1-128.1) | 12 | 0.417 |

**Footnote to Table S3:** abbreviations are as in Table 2 and S1. Chi-square or Fischer's exact test were used to compare categorical variables. Wilcoxon rank sum test was used to compare continuous variables.

**Table S4: Characteristics of non-hospitalized COVID-19 patients.**

| Characteristics | Non-hospitalized COVID-19 patients |
|---|---|
| Number (N) | 26 |
| Age, years | 49.5 (46-62) |
| Sex Male | 38.5% |
| Ethnicity: | |
| - Caucasian | 84.6% |
| - Hispanic | 15.4% |
| - Asian | 0 |
| - African | 0 |
| Co-morbidities: | |
| - Hypertension | 11.5% |
| - CAD | 7.7% |
| - Diabetes | 7.7% |
| - COPD | 0% |
| - CKD | 3.8% |
| - Cancer | 0% |
| - Neurodegenerative disease | 0% |
| N of co-morbidities | |
| - None | 80.8% |
| - 1 | 11.5% |
| - 2 | 3.8% |
| - 3 | 3.8% |
| - 4 | 0 |
| Body Mass Index: | |
| <25 | 31.8% |
| 25-30 | 40.9% |
| >30 | 27.3% |
| Median time (days) from symptoms onset: | |
| - to admission at Emergency Department | 5.5 (3-10) |
| - to first blood sampling for Biobank | 21.5 (5-37) |
| Symptoms at disease onset: | |
| - general | |
| - fever | 68% |
| - headache | 36% |
| - fatigue/malaise | 60% |
| - myalgia/arthralgia | 36% |
| - respiratory | |
| - cough | 60% |
| - dyspnea | 28% |
| - sore throat | 20% |
| - chest pain | 24% |
| - gastrointestinal | |
| - diarrhea | 40% |
| - vomiting/nausea | 12% |
| - abdominal pain | 12% |
| - others | |
| - conjunctivitis | 12% |
| - hypo/anosmia | 28% |
| - hypo/dysgeusia | 40% |
| - skin rash | 1.3% |
| Median time (days) from symptoms to negative RT-PCR swab (95%CI) | 33 (31-35) |
| Median follow up, days (95%CI) | 97 (89-104) |

**Footnote to Table S4:** abbreviations are as in Table 2.

**Supplementary Figures**

**Figure S1**

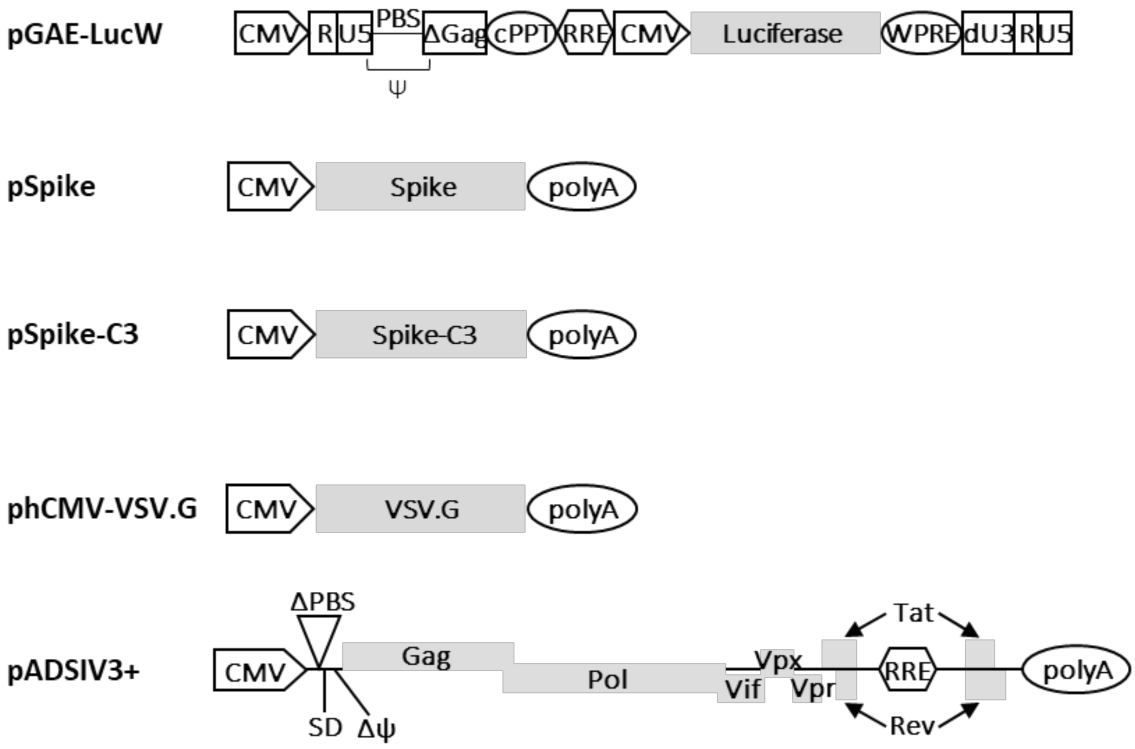


**Footnote to Figure S1: Schematic representation of transfer vectors and plasmids. (A)** Shown are (i) the lentiviral transfer vector plasmid expressing luciferase reporter gene under the control of CMV promoter (pGAE-LucW); (ii) the pseudotyping plasmids expressing full-length Spike (pSpike), cytoplasmic tail truncated Spike (pSpike-C3) and VSV.G envelope (phCMV-VSV.G); (iii) the packaging plasmid providing the proteins for producing the vector particles (pADSIV3+). The packaging signal (ψ), the primer binding site (PBS), the deleted packaging signal (Δψ), the major splice donor (SD), the bovine growth hormone polyadenylation signal (polyA) and the central polypurine tract (cPPT) are indicated.

**Figure S2**

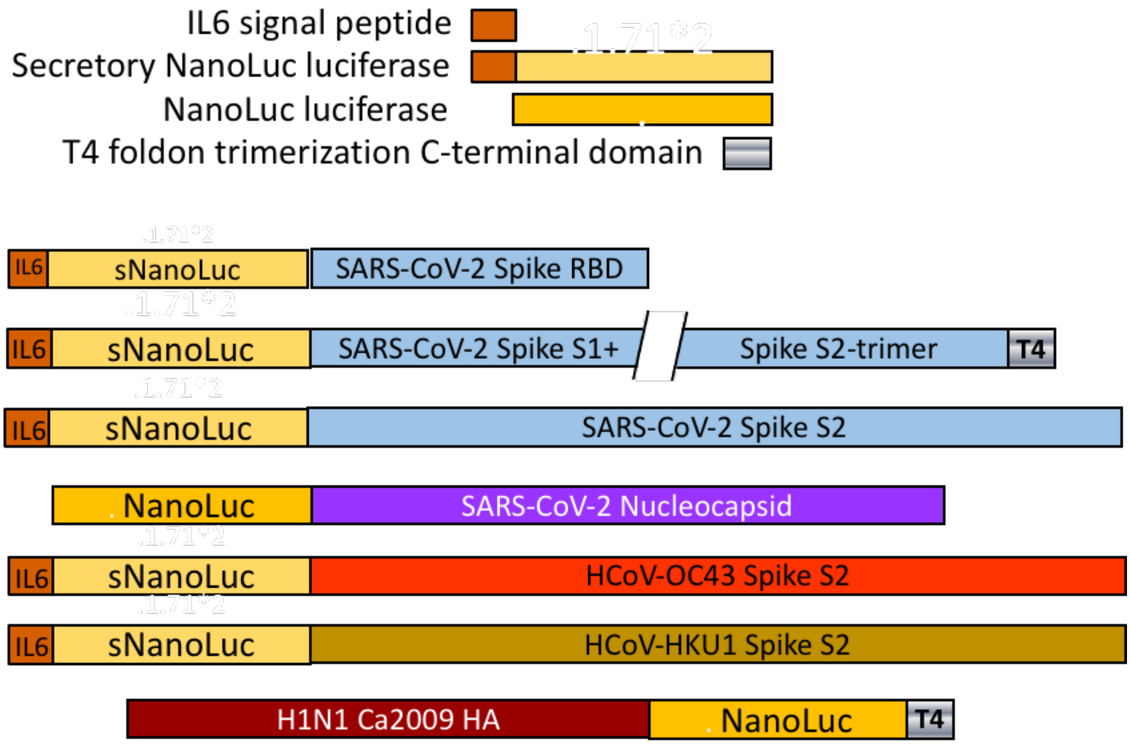


**Footnote to Figure S2. Schematic representation of plasmids used for the LIPS.** Shown are the recombinant nanoluciferase tagged antigens used in this study. See Methods for details on construction.

**Figure S3**

A

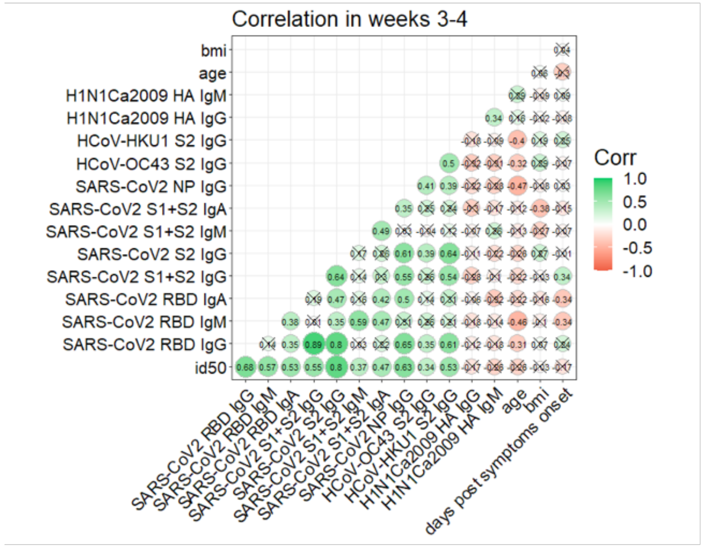
Correlation in weeks 3-4
bmi
age
H1N1Ca2009 HA IgM
H1N1Ca2009 HA IgG
HCoV-HKU1 S2 IgG
HCoV-OC43 S2 IgG
SARS-CoV2 NP IgG
SARS-CoV2 S1+S2 IgA
SARS-CoV2 S1+S2 IgM
SARS-CoV2 S2 IgG
SARS-CoV2 S1+S2 IgG
SARS-CoV2 RBD IgA
SARS-CoV2 RBD IgM
SARS-CoV2 RBD IgG
id50
days post symptoms onset
Corr
1.0
0.5
0.0
-0.5
-1.0

B

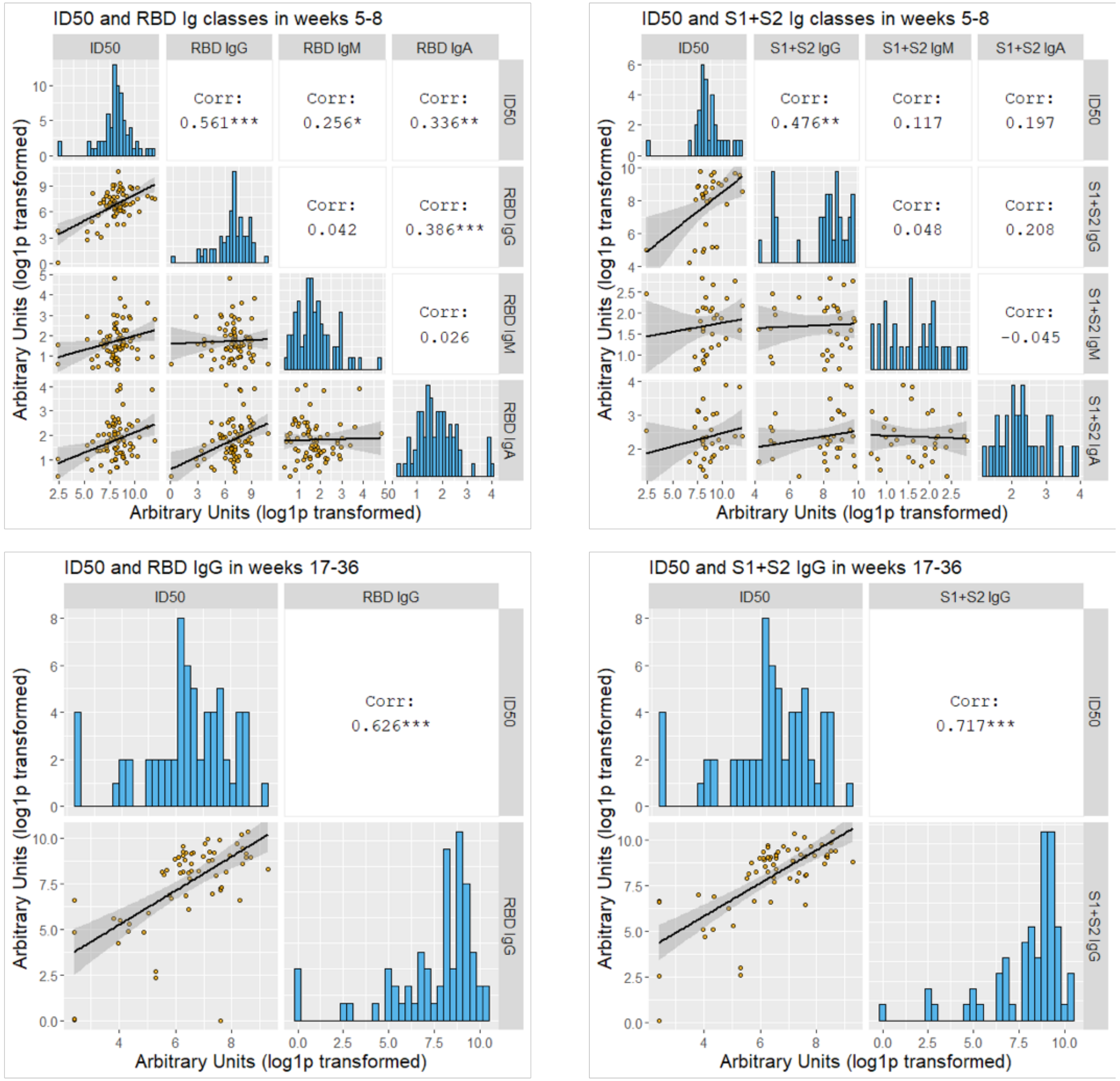
ID50 and RBD Ig classes in weeks 5-8
ID50
RBD IgG
RBD IgM
RBD IgA
Corr: 0.561***
Corr: 0.256*
Corr: 0.336**
Corr: 0.042
Corr: 0.386***
Corr: 0.026
Arbitrary Units (log1p transformed)
ID50 and S1+S2 Ig classes in weeks 5-8
ID50
S1+S2 IgG
S1+S2 IgM
S1+S2 IgA
Corr: 0.476**
Corr: 0.117
Corr: 0.197
Corr: 0.048
Corr: 0.208
Corr: -0.045
Arbitrary Units (log1p transformed)
ID50 and RBD IgG in weeks 17-36
ID50
RBD IgG
Corr: 0.626***
Arbitrary Units (log1p transformed)
ID50 and S1+S2 IgG in weeks 17-36
ID50
S1+S2 IgG
Corr: 0.717***
Arbitrary Units (log1p transformed)

**Footnote to Figure S3: Correlation of anti-SARS-CoV2 spike neutralizing and RBD, S1+S2 antibodies during follow-up of the COVID-19 patients.** Sera of COVID-19 patients, collected at the indicated timepoints from symptoms onset, were measured by LV based-neutralization assay and the LIPS to antigens as indicated in the figures. A) Correlation matrix of the indicated variables in week 3-4. For each pair, the Pearson's correlation coefficient is shown as number and on a color scale. Statistically non-significant correlations are crossed. B) Sera of COVID-19 patients, collected at the indicated timepoints from symptoms onset, were measured by LV based-neutralization assay and the LIPS indicated in grey labels above each row/column. Boxes under the diagonal show each correlation plot of the reciprocal of ID50 and arbitrary units after log10 conversion. Dots correspond to individual measurements, the black line represents the regression line and the grey area its 95%CI. Boxes on the diagonal show as histograms the distribution of values in each assay. Boxes above the diagonal show the corresponding Pearson correlation analysis coefficients.

**Figure S4**

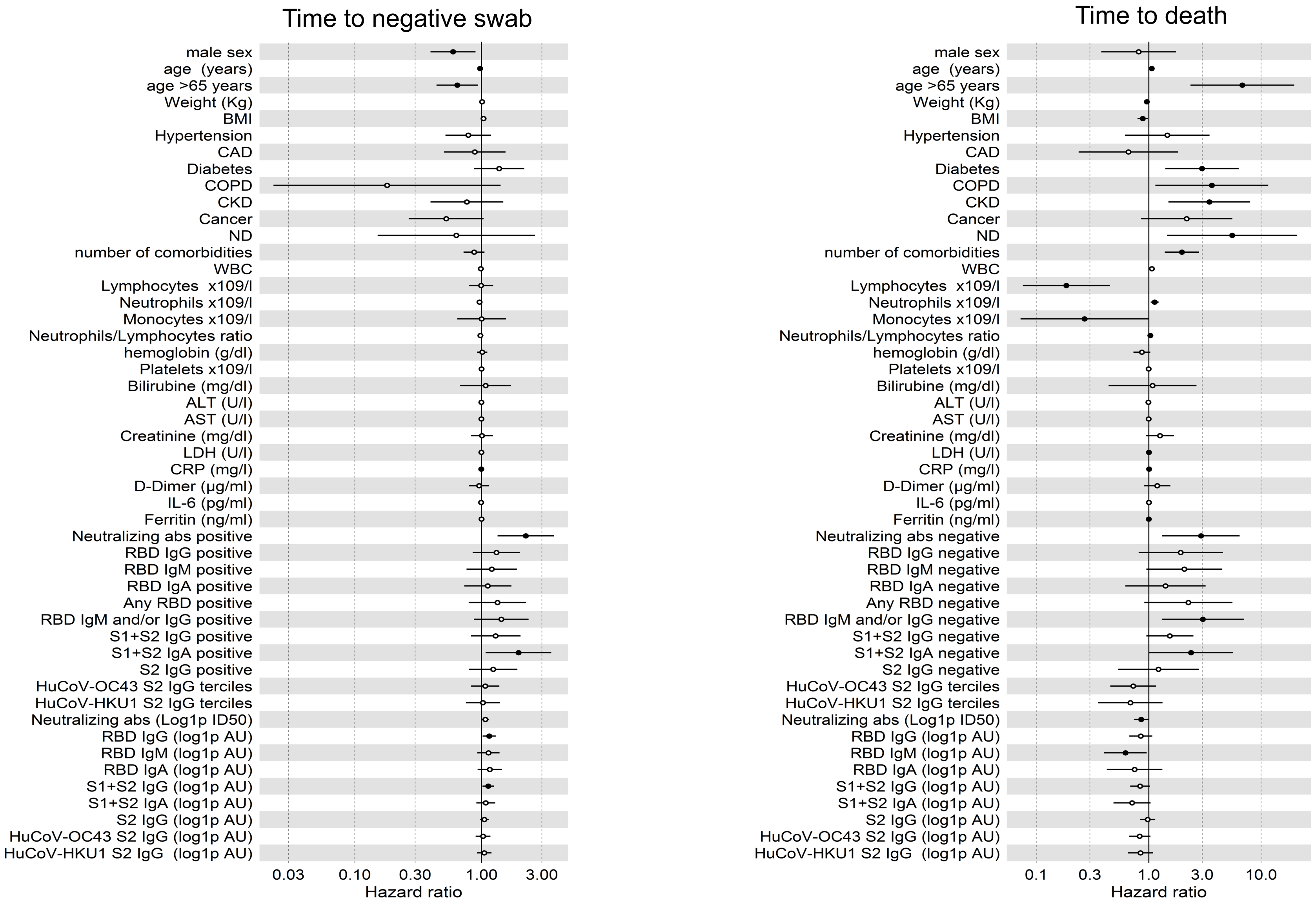

Time to negative swab
male sex
age (years)
age >65 years
Weight (Kg)
BMI
Hypertension
CAD
Diabetes
COPD
CKD
Cancer
ND
number of comorbidities
WBC
Lymphocytes x109/l
Neutrophils x109/l
Monocytes x109/l
Neutrophils/Lymphocytes ratio
hemoglobin (g/dl)
Platelets x109/l
Bilirubine (mg/dl)
ALT (U/l)
AST (U/l)
Creatinine (mg/dl)
LDH (U/l)
CRP (mg/l)
D-Dimer (µg/ml)
IL-6 (pg/ml)
Ferritin (ng/ml)
Neutralizing abs positive
RBD IgG positive
RBD IgM positive
RBD IgA positive
Any RBD positive
RBD IgM and/or IgG positive
S1+S2 IgG positive
S1+S2 IgA positive
S2 IgG positive
HuCoV-OC43 S2 IgG terciles
HuCoV-HKU1 S2 IgG terciles
Neutralizing abs (Log1p ID50)
RBD IgG (log1p AU)
RBD IgM (log1p AU)
RBD IgA (log1p AU)
S1+S2 IgG (log1p AU)
S1+S2 IgA (log1p AU)
S2 IgG (log1p AU)
HuCoV-OC43 S2 IgG (log1p AU)
HuCoV-HKU1 S2 IgG (log1p AU)
0.03
0.10
0.30
1.00
3.00
Hazard ratio
Time to death
male sex
age (years)
age >65 years
Weight (Kg)
BMI
Hypertension
CAD
Diabetes
COPD
CKD
Cancer
ND
number of comorbidities
WBC
Lymphocytes x109/l
Neutrophils x109/l
Monocytes x109/l
Neutrophils/Lymphocytes ratio
hemoglobin (g/dl)
Platelets x109/l
Bilirubine (mg/dl)
ALT (U/l)
AST (U/l)
Creatinine (mg/dl)
LDH (U/l)
CRP (mg/l)
D-Dimer (µg/ml)
IL-6 (pg/ml)
Ferritin (ng/ml)
Neutralizing abs negative
RBD IgG negative
RBD IgM negative
RBD IgA negative
Any RBD negative
RBD IgM and/or IgG negative
S1+S2 IgG negative
S1+S2 IgA negative
S2 IgG negative
HuCoV-OC43 S2 IgG terciles
HuCoV-HKU1 S2 IgG terciles
Neutralizing abs (Log1p ID50)
RBD IgG (log1p AU)
RBD IgM (log1p AU)
RBD IgA (log1p AU)
S1+S2 IgG (log1p AU)
S1+S2 IgA (log1p AU)
S2 IgG (log1p AU)
HuCoV-OC43 S2 IgG (log1p AU)
HuCoV-HKU1 S2 IgG (log1p AU)
0.1
0.3
1.0
3.0
10.0
Hazard ratio

**Footnote to Figure S4: Univariate Hazard Ratios (HR) for time to a negative SARS-CoV-2 viral RNA RT-PCR of the nasopharyngeal swab and to death of COVID-19 patients.** Forest plots of Hazard Ratios obtained by univariable Cox regression analysis of the shown variables at the time of the in-hospital serum sampling. The analysis was adjusted for sex and age. Dots represent the HR, filled dots stand for $p<0.05$.

**Figure S5**

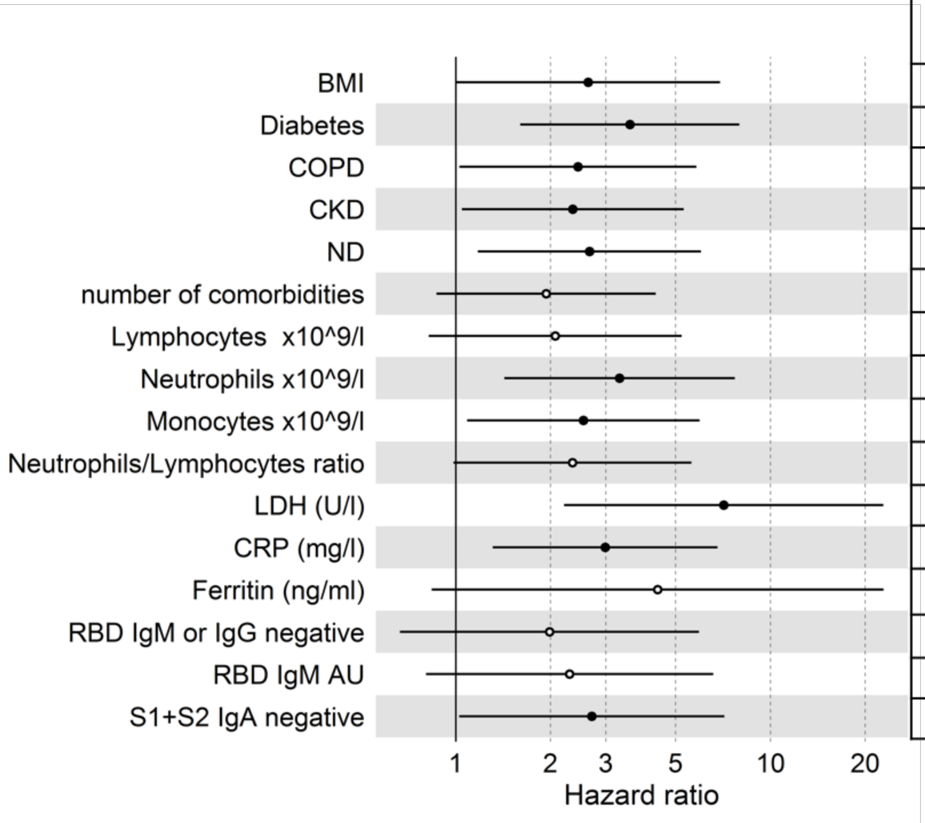


| Significance | HR | 95·0%CI | |
|---|---|---|---|
| | | Lower | Upper |
| 0·049 | 2·635 | 1·004 | 6·918 |
| 0·002 | 3·575 | 1·604 | 7·966 |
| 0·044 | 2·445 | 1·026 | 5·826 |
| 0·039 | 2·354 | 1·046 | 5·302 |
| 0·019 | 2·659 | 1·176 | 6·015 |
| 0·107 | 1·937 | 0·868 | 4·322 |
| 0·123 | 2·071 | 0·821 | 5·225 |
| 0·005 | 3·316 | 1·428 | 7·703 |
| 0·031 | 2·546 | 1·087 | 5·962 |
| 0·055 | 2·350 | 0·984 | 5·614 |
| 0·001 | 7·104 | 2·208 | 22·855 |
| 0·009 | 2·984 | 1·311 | 6·794 |
| 0·080 | 4·382 | 0·839 | 22·895 |
| 0·219 | 1·986 | 0·665 | 5·933 |
| 0·120 | 2·300 | 0·804 | 6·578 |
| 0·044 | 2·707 | 1·026 | 7·142 |

**Footnote to Figure S5: Bivariate Hazard Ratios (HR) for time to death of COVID-19 patients.** Forest plot of Hazard Ratios (HR) for neutralizing antibodies and survival in COVID-19 patients calculated with multivariable Cox regression analysis. The analysis used a neutralization negative score, corrected for age and sex, and the shown variables measured at the time of in-hospital serum sampling. Dots represent the HR, filled dots stand for $p<0.05$. Abbreviations are as in Table 1; Coronary Artery Diseases (CAD), Chronic obstructive pulmonary disease (COPD), Chronic Kidney Disease (CKD), Neurodegenerative disease (ND).

**Figure S6**

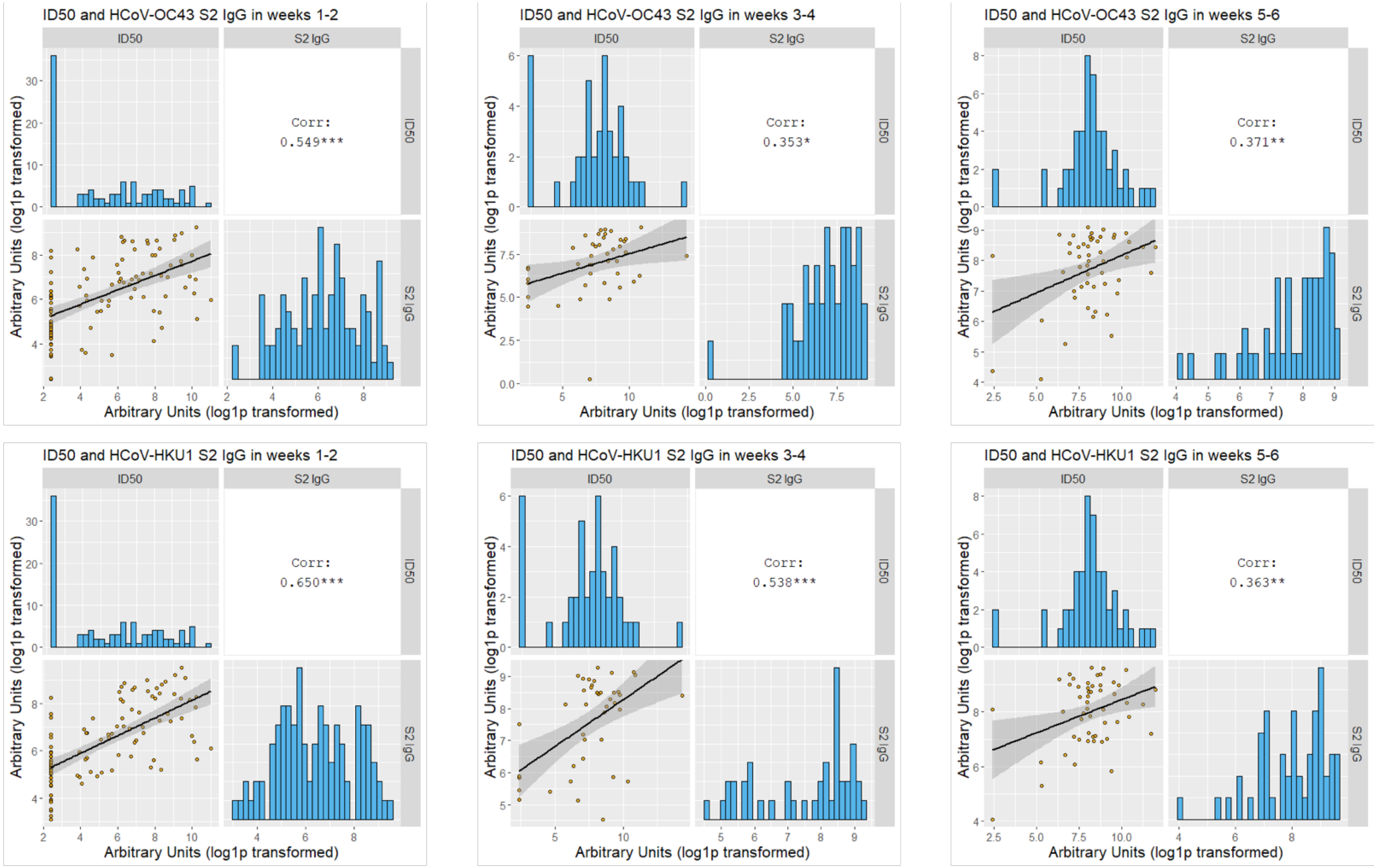
ID50 and HCoV-OC43 S2 IgG in weeks 1-2
ID50
S2 IgG
Corr:
0.549***
Arbitrary Units (log1p transformed)
Arbitrary Units (log1p transformed)
ID50 and HCoV-OC43 S2 IgG in weeks 3-4
ID50
S2 IgG
Corr:
0.353*
Arbitrary Units (log1p transformed)
Arbitrary Units (log1p transformed)
ID50 and HCoV-OC43 S2 IgG in weeks 5-6
ID50
S2 IgG
Corr:
0.371**
Arbitrary Units (log1p transformed)
Arbitrary Units (log1p transformed)
ID50 and HCoV-HKU1 S2 IgG in weeks 1-2
ID50
S2 IgG
Corr:
0.650***
Arbitrary Units (log1p transformed)
Arbitrary Units (log1p transformed)
ID50 and HCoV-HKU1 S2 IgG in weeks 3-4
ID50
S2 IgG
Corr:
0.538***
Arbitrary Units (log1p transformed)
Arbitrary Units (log1p transformed)
ID50 and HCoV-HKU1 S2 IgG in weeks 5-6
ID50
S2 IgG
Corr:
0.363**
Arbitrary Units (log1p transformed)
Arbitrary Units (log1p transformed)

**Footnote to Figure S6: HCoV-HKU1 and -OC43 S2 IgG correlation with neutralization is temporary.** Sera of COVID-19 patients, collected at the indicated timepoints from symptoms onset, were measured by LV based-neutralization assay and the LIPS indicated in grey labels above each row/column. Boxes under the diagonal show each correlation plot of the reciprocal of ID50 and arbitrary units after log10 conversion. Dots correspond to individual measurements, the black line represents the regression line and the grey area its 95%CI. Boxes on the diagonal show as histograms the distribution of values in each assay. Boxes above the diagonal show the corresponding Pearson correlation analysis coefficients.

**Figure S7**

A

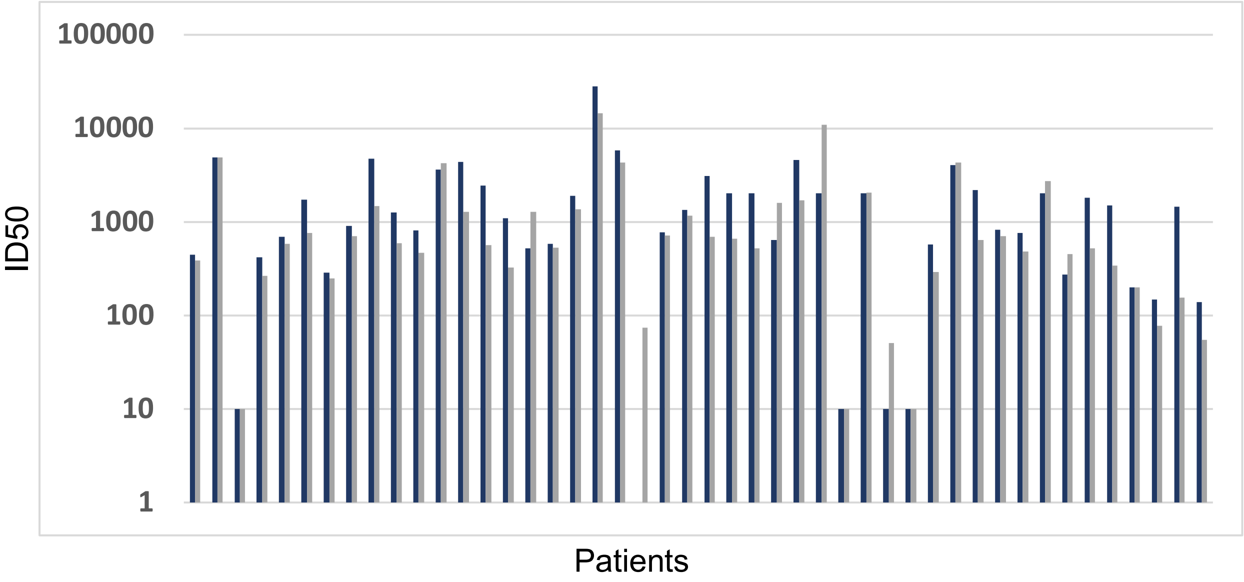


B

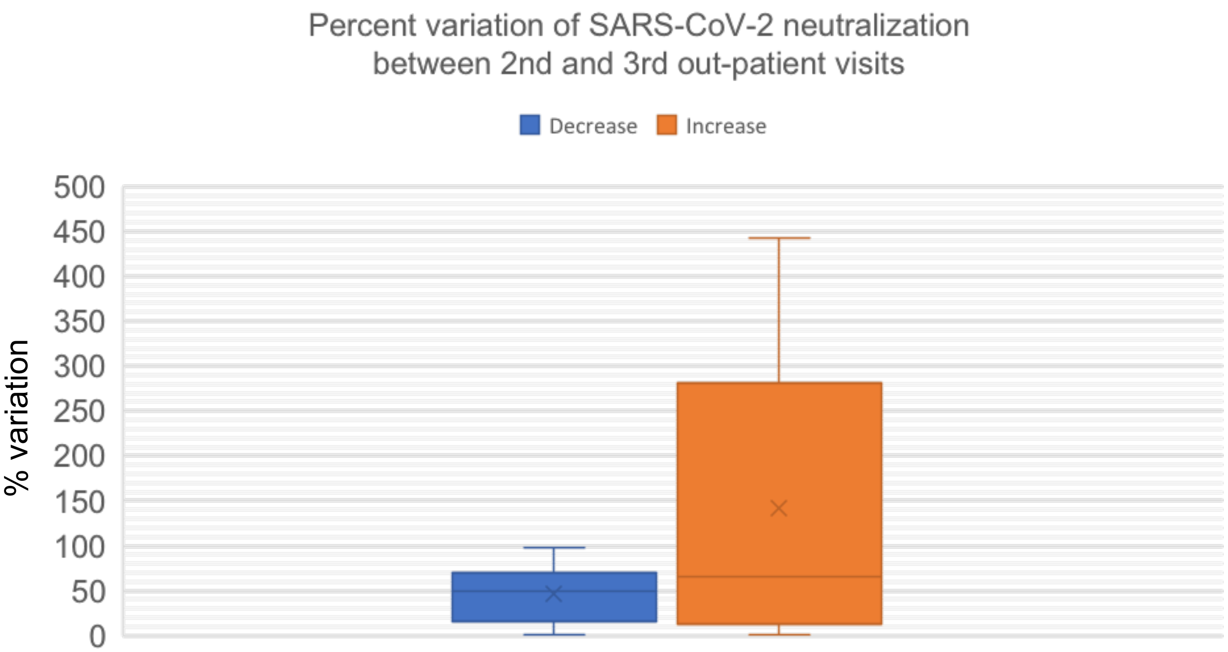


| | | |
|---|---|---|
| N individuals | 34 | 9 |
| Median % variation | 48·78 | 65·90 |
| Range (min-max) | 0·3 - 98·13 | 1·29 - 441·92 |
| N with variation >30% | 22 | 6 |

**Footnote to Figure S7: Variations of the SARS-CoV-2 neutralizing titers between the $2^{nd}$ and $3^{rd}$ out-patient visit.** Indicated is**:** (A) the ID50 titer at the $2^{nd}$ and $3^{rd}$ visit (dark and light grey bars, respectively) of each of the 43 patients tested for nAbs. (B) the percent variation observed. The $3^{rd}$ visit occurred between September 22nd and November 6th 2020, when the Rt in the same geographical area of the San Raffaele Hospital, the Lombardy region, ramped-up from 0.86 (CI: 0.73-1.01) at the end of September to 1.61 (CI: 1.08-2.33) in the first week of November 2020. (http://www.salute.gov.it/portale/nuovocoronavirus/dettaglioNotizieNuovoCoronavirus.jsp?lingua=italiano&menu=notizie&p=dalministero&id=5093).

**Figure S8**

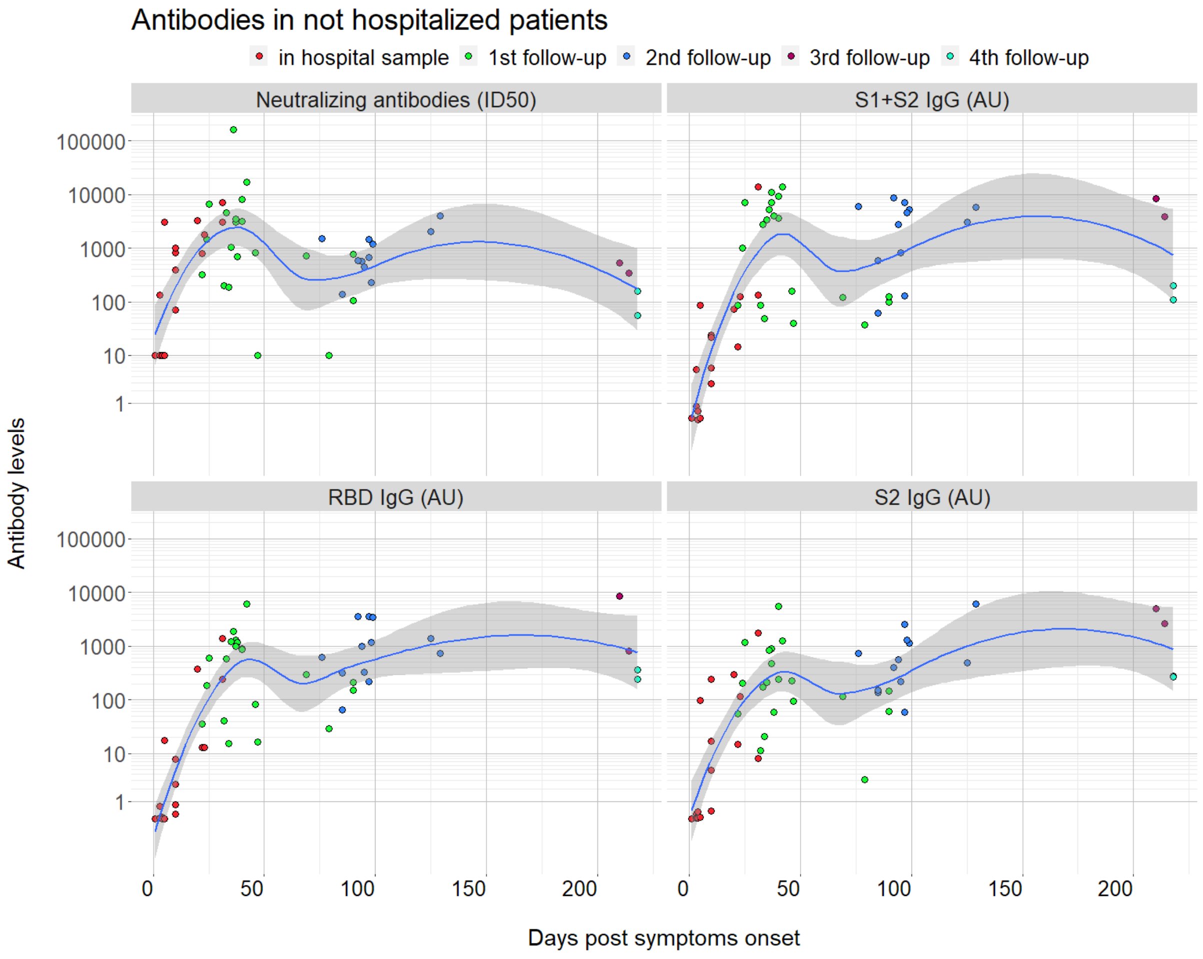


**Footnote to Figure S8: Development and kinetics of SARS-CoV-2 antibody responses of non-hospitalized COVID-19 patients.** Each colored dot corresponds to the reciprocal of the ID50 of a serum of a given infected individual. Sampling occurred during hospital attendance (ER or ward), and at follow-up visits post discharge; colors define the number of the visits attended. In Table 1 the serum sample availability of the non-hospitalized COVID-19 patients is described. The clinical characteristics are described in Table S4.
